# Pathogenicity Reassessment and Novel Variant Discovery in Inherited Retinal Disease through Population-Scale Genomics in the United Arab Emirates

**DOI:** 10.1101/2025.10.14.25337784

**Authors:** Budour Alkaf, Wadha Mohammed Abdulrahman, Sahar Al Marzooqi, Daniel Sanchez, Andreas Henschel, Aashish Jha, Abdelrahman Saad, Ahmed Al Awadhi, Albarah El-Khani, Aleksander Medvedev, Amna Alsuwaidi, Antonio Milano, Asma Al Mannaei, Ayesha Al Ali, Azza Attia, Aziz Khan, Eduardo Beltrame, Fahed Al Marzooqi, Gurunath Katagi, Haiguo Wu, Hanil Sajad, Imam Chishty, Islam Eltantawy, Joseph Mafofo, Judith Arres, Kristine Wong, Lawrence Petalidis, Maaz Shaikh, Marie Ibrahim, Mohamed El-Hadidi, Omar Soliman, Pierre Zalloua, Pradhuman Gupta, Raony Cardenas, Sana Islam, Shalini Behl, Shobith Pejathaya, Shreyaa Chandrashekar, Thyago Cardoso, Val Zvereff, Vinay Kusuma, Vineet Kumar, Youssef Idaghdour, Mohamed Alameri, Javier Quilez, Arif Khan, Tiago Magalhaes

## Abstract

**Background:** Pathogenicity classifications for rare disease variants are largely derived from clinically ascertained cohorts, in which penetrance is often assumed to be near-complete. As genomic sequencing expands into population-scale screening, particularly in underrepresented populations, both the penetrance of known variants and the relevance of novel or population-enriched variants require systematic evaluation. Inherited retinal diseases (IRDs), genetically heterogeneous and clinically variable, represent an ideal setting in which to assess how ascertainment-based pathogenicity assignments translate to real-world population risk.

**Methods:** We analyzed whole-genome sequencing data from 504,000 participants in the Emirati Genome Program, including 426,382 individuals with linked longitudinal electronic health records and 75,184 genetically reconstructed family units. IRD phenotypes were defined through a phenome-wide association approach to derive enriched retinal ICD-10 code sets. Variants in 339 IRD-associated genes were assessed using a population-based framework integrating enrichment testing, penetrance tiering, and family-based concordance across pathogenic or likely pathogenic variants (P/LP), variants of uncertain significance (VUS), and novel variants not previously classified in clinical variant databases, yielding a population-calibrated IRD panel.

**Results:** Among 24,100 variants observed in individuals with IRD-compatible genotypes (259 P/LP, 11,888 VUS, and 11,953 novel variants), 533 variants were prioritized. P/LP variants showed the highest proportional enrichment (15.4% vs. 3.4% of VUS and 3.2% of novel variants), with the strongest population-level signals being P/LP variants in *KCNV2* and *ABCA4*, both markedly enriched in the Emirati cohort relative to global reference data and consistent with their predominance among affected individuals at regional ophthalmology clinics. Restricting prioritization to P/LP variants alone would have captured only 37 variants; inclusion of VUS and novel variants expanded the panel more than 14-fold. Conversely, 9.3% of P/LP variants lacked population-level enrichment, including 3.1% with null penetrance despite adequate carrier representation. Family-based concordance supported 404 of 533 prioritized variants, defining the population-calibrated panel; 91.8% of these were VUS or novel.

**Conclusions:** Penetrance of IRD-associated variants is heterogeneous and frequently lower than inferred from clinical cohorts. Population-scale calibration identified clinically relevant VUS and novel variants, including population-specific risk alleles absent from global databases, while revealing limited real-world support for a subset of established P/LP classifications. These findings underscore that assertion-based pathogenicity classifications require recalibration against real-world disease expression before deployment in population-scale genomic screening, particularly in underrepresented populations.

## Introduction

As genome sequencing is increasingly deployed beyond specialty clinics into population-scale screening initiatives, interpreting the clinical relevance of genetic variants remains a central challenge^1, 2^. Pathogenicity classifications are largely derived from clinically ascertained cohorts in which affected individuals are preferentially evaluated, and penetrance is often implicitly assumed to be high^3, 4, 5^. In addition, clinical interpretation relies heavily on pathogenic variants identified in well-studied populations, and extrapolating these classifications to underrepresented groups may overlook population-specific disease signals not captured in existing databases. When such categorical classifications are applied to unselected populations, however, real-world disease expression may differ substantially^6, 7, 8^. Recent population-scale studies have demonstrated that many variants associated with Mendelian conditions exhibit lower penetrance than historically inferred^6, 8^, underscoring the need to recalibrate pathogenicity in the context of population-scale deployment.

IRDs represent a clear example of these challenges. They comprise a genetically and clinically heterogeneous group of predominantly monogenic disorders^9^, and represent a leading cause of vision loss worldwide, with an estimated prevalence of approximately 1 in 3,000^10^. More than 330 genes have been implicated^11^, encompassing diverse biological pathways and inheritance patterns. Beyond genetic heterogeneity, IRDs also exhibit marked clinical variability in age at onset, severity, progression, and syndromic involvement, further complicating accurate diagnosis and gene-based stratification^12, 13, 14^.

In the Arabian Gulf, the burden of inherited ocular disease is reported at substantially higher frequencies than global IRD prevalence estimates, with regional reports suggesting that up to 5% of the population may be affected by genetic conditions involving the eye^15, 16^. Although population-based IRD prevalence data specific to the region remain limited, elevated consanguinity is expected to increase autosomal recessive disease through founder effects and genetic drift^17, 18, 19^, potentially enriching population-specific variants that may be underrepresented in global reference databases.

The Emirati Genome Program (EGP), one of the world’s largest population-scale whole-genome sequencing (WGS) initiatives, offers a unique opportunity to evaluate genotype–phenotype relationships at national scale. The program integrates deep WGS of more than half a million individuals with longitudinal electronic health records (EHR), derived from the Abu Dhabi health information exchange (Malaffi), together with extensive multigenerational family structures characteristic of the Emirati population. This integrated resource enables population-based penetrance estimation and inheritance-aware evaluation of disease-associated variants.

Here, we report the application of a population-scale genomic framework to first determine the prevalence of genetic variants known to be associated with IRD and uncover previously unreported IRD-associated variants which are likely to be private of the Emirati population. By integrating genomics and longitudinal clinical data, we developed and applied a scalable analytic framework that allowed us to construct a population-calibrated IRD variant panel as well as to estimate the penetrance of its variants and prioritize them for clinical interpretation. In addition, we use family relationships at population-scale to support our findings and extend variant prioritization at the familial level. Through this approach, we provide a framework for evaluating pathogenicity across the genotype–phenotype spectrum, with implications for clinical interpretation and genomic screening in IRDs and other rare diseases.

## Methods

### Ethics Approval and Consent

Genomic sequencing and participant consent procedures were conducted under the governance of the EGP. The study was approved by the Department of Health–Abu Dhabi Human Research Ethics Committee (IRB protocol DOH/ADHRTC/2025/1032). All participants provided written informed consent for the use of de-identified genomic and clinical data.

### Study Population and Family Reconstruction

We used WGS datasets for 504,000 EGP participants (Data Freeze Version 1). Longitudinal electronic health record (EHR) data were extracted through Malaffi for EGP participants (phenoEGP), capturing clinical encounters from 2013 to 2024. The integrated EGP–phenoEGP cohort comprised 426,382 participants with both WGS and linked EHR data.

Using WGS-derived genotypes for the genetic markers profiled in the Illumina Global Screening Array (GSA)^20^, kinship analysis was performed across 504,000 individuals in the EGP. Pairwise biological relationships were inferred using KING^21^, and multigenerational family networks were reconstructed using parent–offspring relationships and age information, yielding 75,184 family units. Family units were defined as clusters anchored by the earliest inferred generation and including direct descendants, enabling inheritance-aware variant interpretation and family-based analyses.

### IRD Variant Panel Construction

We curated three sets of variants within genes associated with IRD, as defined by the RetNet database (339 genes; accessed April 2025). The first set comprised ClinVar-classified^22^ pathogenic or likely pathogenic (P/LP) variants with at least one review star (ClinVar VCF release November 2024), supplemented by 50 well-established pathogenic variants identified through clinical evaluation of affected Emirati families at Cleveland Clinic Abu Dhabi (P/LP; N = 15,795). The second set included ClinVar-reported variants of uncertain significance (VUS; N = 132,445). The third set consisted of variants not present in ClinVar^22^, predicted to have moderate or high functional impact by the Variant Effect Predictor^23^ (VEP, Ensembl), and observed at a minor allele frequency (MAF) below 1% in the EGP cohort (N = 48,366). These variants were evaluated as previously unclassified candidates for IRD association.

### Identification of Core Retinal Phenotypes

To define retinal phenotypes suitable for population-scale penetrance analysis, we performed a phenome-wide association study (PheWAS) across all low-frequency ICD-10 diagnoses (<1% prevalence) in the EGP–phenoEGP cohort, anchored to carriers of known IRD-associated P/LP variants (**Supplementary Appendix, ST1, ST2**). This approach addressed the clinical heterogeneity of IRDs^3^ and the limitations of relying on single diagnostic codes in real-world electronic health records^6^.

The PheWAS identified five ICD-10 subchapters (H35, H50, H53, H54, and H55) that were consistently enriched among variant carriers and collectively defined a set of core IRD-related phenotypes (“Top5”) (**Supplementary Appendix, SF1**). Individuals with at least one recorded Top5 phenotype were classified as IRD cases for all population-level analyses.

### Population-Based Variant Prioritization Framework

We developed a structured population-based variant prioritization framework integrating enrichment testing, penetrance estimation, and family-based validation (**Supplementary Appendix, SF2**). Each variant was tested for enrichment among IRD cases relative to non-carriers using Fisher’s exact test. Variants demonstrating a positive risk difference (RD > 0) and nominal evidence of enrichment (P < 0.05) were carried forward for penetrance estimation.

Variant-level penetrance was defined as the proportion of individuals carrying IRD-associated genotypes who were classified as IRD cases. For loci in genes annotated as autosomal dominant for IRD, both heterozygous and homozygous variant carriers were considered to have IRD-compatible genotypes. For loci annotated with other modes of inheritance, only homozygous variant carriers were classified as having IRD-compatible genotypes. Variants were assigned to penetrance tiers based on observed penetrance: Tier 1 (100%), Tier 2 (≥50% to <100%), Tier 3 (≥25% to <50%), Tier 4 (≥15% to <25%), and Tier 5 (>0% to <15%). Prioritized variants were defined as those assigned to Tiers 1–3, corresponding to moderate to complete penetrance.

Variants without statistically significant enrichment (RD ≤ 0 or P ≥ 0.05) were not assigned to penetrance tiers. Among these, variants observed in at least 10 individuals with IRD-compatible genotypes were denoted separately to indicate stable population-level estimates. Variants with ≥10 genotype-compatible carriers and no documented IRD cases (penetrance = 0) were designated as having null penetrance and evaluated separately (Supplementary Appendix, ST3).

To assess consistency between population-prioritized variants and segregation patterns, prioritized variants were further evaluated using a family-based concordance analysis across reconstructed family units (**Supplementary Appendix, SF3)**. For each variant, we quantified the number of family units in which at least one IRD case carried the variant and evaluated whether all IRD cases within a family were variant carriers. Within the remaining family units, when multiple prioritized variants were present, the variant with the strongest population-level support was designated as the best-explaining variant and considered concordant.

## Results

### Population-Scale Landscape of IRD-associated Genetic Variation

All analyses were performed in 426,382 participants of the EGP-phenoEGP cohort. Among individuals with IRD-compatible genotypes, we identified 24,100 variants observed in at least one IRD genotype carrier across 330 IRD-associated genes (**Supplementary Appendix, ST4).** These variants comprised 259 P/LP, 11,888 VUS and 11,953 novel variants (not previously classified in clinical variant databases, **Methods**). Relative to the total screened variant sets, these represent 1.6% of P/LP variants, 9.0% of VUS, and 24.7% of novel variants that were observed in IRD genotype carriers. Together, these results define the population-scale landscape of IRD-associated genetic variation and provide the basis for a scalable framework for population-calibrated IRD variant prioritization.

### Building a Population-Calibrated IRD Variant Panel

IRD variants observed in the EGP-phenoEGP cohort were evaluated using a population-based variant prioritization framework (Methods). Variants were first assessed for enrichment among IRD cases, followed by estimation of observed penetrance, defined as the proportion of individuals with IRD-compatible genotypes who had an IRD diagnosis. Variants meeting these criteria were then assigned to penetrance tiers and designated as prioritized variants based on tier classification.

A total of 41,725 individuals (9.8%) of the EGP–phenoEGP cohort were classified as IRD cases. P/LP variants showed the highest proportional enrichment, with 40 of 259 variants (15.4%) demonstrating significant association with IRD phenotypes (risk difference (RD) > 0, P < 0.05). However, restricting analyses to P/LP variants captured only a small fraction of the overall enrichment signal. Expanding the analysis to include VUS increased the number of enriched variants more than tenfold (407 of 11,888 [3.4%]), and inclusion of novel variants identified an additional 381 enriched variants (3.2%) (**Table1**). Notably, the identification of enriched variants among previously unclassified candidates underscores the extent to which clinically relevant disease signal remains undetected when variant interpretation relies solely on existing pathogenicity databases, particularly in historically understudied populations.

The remaining 23,272 variants (97%) did not show significant enrichment and were not interpreted as evidence for or against disease association. Within this non-enriched group, 24 of 259 P/LP variants (9.3%) were observed in at least 10 individuals with IRD-compatible genotypes but failed to demonstrate significant enrichment. Among these, 8 P/LP variants (3.1% of all P/LP variants assessed) exhibited null penetrance, with no documented IRD cases despite adequate carrier representation (**Table 1**). These findings provide population-level evidence that a subset of variants currently classified as P/LP may warrant reassessment in population-based screening contexts.

**Table 1.** Population-calibrated stratification of inherited retinal disease variants by penetrance tier across individuals, families, and genes. This table summarizes variants demonstrating significant population-level enrichment for IRD, their distribution across penetrance tiers, and counts supported by familial concordance. Counts are shown across individuals, IRD cases, family units, genes, and variants under progressively inclusive panel definitions (P/LP; P/LP + VUS; P/LP + VUS + Novel).

|  | Panel | Tier 1<br>(Pen=1.0) | Tier 1–2<br>(≥0.50) | Tier 1–3<br>(≥0.25) | Tier 1–4<br>(≥0.15) | Tier 1–5<br>(>0) | Not enriched<br>(overall) | Not enriched<br>(≥10 carriers) | Null<br>penetrance |
| --- | --- | --- | --- | --- | --- | --- | --- | --- | --- |
| Population-level Enrichment |  |  |  |  |  |  |  |  |  |
| Individuals (N=426,382) | P/LP | 45 | 197 | **300** | 667 | 1,681 | 957 | **612** | **106** |
|  | P/LP + VUS | 103 | 606 | **2,771** | 13,668 | 95,794 | 291,612 | 285,257 | 6,622 |
|  | P/LP + VUS + Novel | 183 | 1,090 | **5,157** | 21,795 | 131,415 | 366,526 | 360,806 | 13,147 |
| IRD cases (N=41,725) | P/LP | 45 | 155 | **191** | 253 | 378 | 127 | **49** | — |
|  | P/LP + VUS | 103 | 425 | **1,079** | 2,926 | 11,930 | 27,811 | 27,173 | — |
|  | P/LP + VUS + Novel | 183 | 758 | **1,974** | 4,750 | 16,422 | 35,321 | 34,766 | — |
| Family units (N=75,184) | P/LP | 30 | 138 | **199** | 429 | 1,010 | 575 | **344** | **54** |
|  | P/LP + VUS | 68 | 387 | **1,508** | 6,297 | 29,191 | 67,879 | 67,072 | 3,075 |
|  | P/LP + VUS + Novel | 117 | 667 | **2,639** | 9,788 | 38,831 | 73,338 | 72,958 | 5,756 |
| Family units with IRD case<br>(N=24,499) | P/LP | 30 | 116 | **148** | 191 | 296 | 107 | **42** | — |
|  | P/LP + VUS | 68 | 302 | **782** | 2,070 | 8,009 | 18,035 | 17,705 | — |
|  | P/LP + VUS + Novel | 117 | 525 | **1,381** | 3,287 | 10,763 | 21,779 | 21,525 | — |
| Genes (N=330) | P/LP | 13 | 23 | **26** | 27 | 27 | 84 | **10** | **7** |
|  | P/LP + VUS | 40 | 91 | **130** | 153 | 157 | 316 | 274 | 156 |
|  | P/LP + VUS + Novel | 64 | 153 | **194** | 215 | 217 | 327 | 307 | 199 |
| Variants (N=24,100) | P/LP | 17 | 31 | **37** | 39 | 40 | 219 | **24** | **8** |
|  | P/LP + VUS | 46 | 142 | **266** | 387 | 447 | 11,700 | 3,613 | 442 |
|  | P/LP + VUS + Novel | 83 | 288 | **533** | 730 | 828 | 23,272 | 6,851 | 904 |
| Familial Validation |  |  |  |  |  |  |  |  |  |
| Individuals (N=426,382) | P/LP | 37 | 189 | **292** | 659 | 1,673 | — | — | — |
|  | P/LP + VUS | 70 | 517 | **2,567** | 13,463 | 95,631 | — | — | — |
|  | P/LP + VUS + Novel | 117 | 823 | **4,605** | 21,115 | 130,184 | — | — | — |
| IRD cases (N=41,725) | P/LP | 37 | 147 | **183** | 245 | 370 | — | — | — |
|  | P/LP + VUS | 70 | 356 | **977** | 2,836 | 11,865 | — | — | — |
|  | P/LP + VUS + Novel | 117 | 569 | **1,699** | 4,485 | 16,145 | — | — | — |
| Family units (N=75,184) | P/LP | 19 | 104 | **153** | 343 | 840 | — | — | — |
|  | P/LP + VUS | 45 | 298 | **1,265** | 5,724 | 30,942 | — | — | — |
|  | P/LP + VUS + Novel | 76 | 467 | **2,159** | 8,927 | 41,576 | — | — | — |
| Family units with IRD case<br>(N=24,499) | P/LP | 19 | 86 | **111** | 147 | 235 | — | — | — |
|  | P/LP + VUS | 45 | 229 | **626** | 1,764 | 7,230 | — | — | — |
|  | P/LP + VUS + Novel | 76 | 368 | **1,068** | 2,784 | 9,732 | — | — | — |
| Genes (N=330) | P/LP | 10 | 20 | **23** | 23 | 23 | — | — | — |
|  | P/LP + VUS | 26 | 75 | **118** | 118 | 118 | — | — | — |
|  | P/LP + VUS + Novel | 42 | 117 | **168** | 168 | 168 | — | — | — |
| Variants (N=24,100) | P/LP | 13 | 27 | **33** | 35 | 36 | — | — | — |
|  | P/LP + VUS | 30 | 107 | **219** | 335 | 393 | — | — | — |
|  | P/LP + VUS + Novel | 49 | 191 | **404** | 589 | 679 | — | — | — |

Penetrance (Pen): proportion of individuals with IRD-compatible genotypes with a documented Top5 retinal diagnosis. Counts are cumulative: P/LP = pathogenic/likely pathogenic variants; P/LP+VUS adds variants of uncertain significance; P/LP+VUS+Novel adds previously unclassified variants absent in clinical variant databases. Tier 1–3= prioritized population-calibrated panel (penetrance ≥25%). Not enriched (≥10 carriers, amber): P/LP variants with ≥10 carriers lacking significant enrichment. Null penetrance (≥10 carriers, red): P/LP variants with ≥10 carriers and no documented IRD cases. — indicates not applicable.

Variant scoring by composite enrichment (RD x –log_10_P) highlighted the strongest population-level signals to be a P/LP variant in *KCNV2* and another P/LP in *ABCA4* (**Supplementary Appendix, ST4)**. Notably, both variants are represented in IRD diagnostic panels currently used in clinical practice at regional ophthalmology clinics in the UAE^24^. Neither variant was observed in gnomAD genome sequences, and both were present at substantially higher frequencies in the EGP than in gnomAD exome data (*KCNV2*: EGP AF = 8.1×10⁻⁴ vs. gnomAD exome AF = 5.6×10⁻⁶; *ABCA4*: EGP AF = 2.3×10⁻³ vs. gnomAD exome AF = 6.2×10⁻⁷), consistent with population-specific enrichment in the Emirati population.

Enriched variants were subsequently evaluated for penetrance and assigned to penetrance tiers, resulting in a total of 533 prioritized variants (Tiers 1–3), including 37 P/LP variants, 229 VUS, and 267 novel variants (**Table 1; Supplementary Appendix, SF4**). Notably, only 7% of prioritized variants were P/LP, indicating that population-level penetrance calibration substantially expands beyond conventional pathogenic classifications. When analyses were expanded to include possible IRD-compatible genotypes (heterozygous carriers in genes with uncertain inheritance; **Supplementary Methods**), the number of prioritized variants increased to 992 (**Supplementary Appendix, ST6**), with 31 additional Tier 1 variants reaching complete observed penetrance, suggesting dominant inheritance behavior among these newly prioritized variants.

To account for the progressive onset of many IRDs, we applied the population-based variant prioritization framework in individuals aged ≥35 years. This age-stratified analysis identified 273 variants that reached Tier 1–3 penetrance in the ≥35-year subset but did not meet prioritization criteria in the full cohort. These variants were concentrated in genes associated with retinitis pigmentosa, one of the most common IRDs characterized by slow disease progression^25^, including *VWAB*, *SNRNP200*, and *PRPF8*.

To assess whether variants prioritized using the broader Top5 retinal phenotype captured disease signal under a stricter clinical definition, we conducted an independent enrichment analysis using a curated set of IRD-specific ICD-10 codes derived from an external population-based study^6^. In the P/LP variant group, enrichment using the broader Top5 phenotype (OR 2.2) was comparable to effect sizes reported in the external study using a curated retinal code set^6^. Under the stricter IRD-specific phenotype definition, enrichment increased substantially (OR 20.6) (**Figure 1; Supplementary Methods**), consistent with the marked increase observed in the external study, despite differences in cohort composition and variant inclusion criteria.

**Figure 1.**
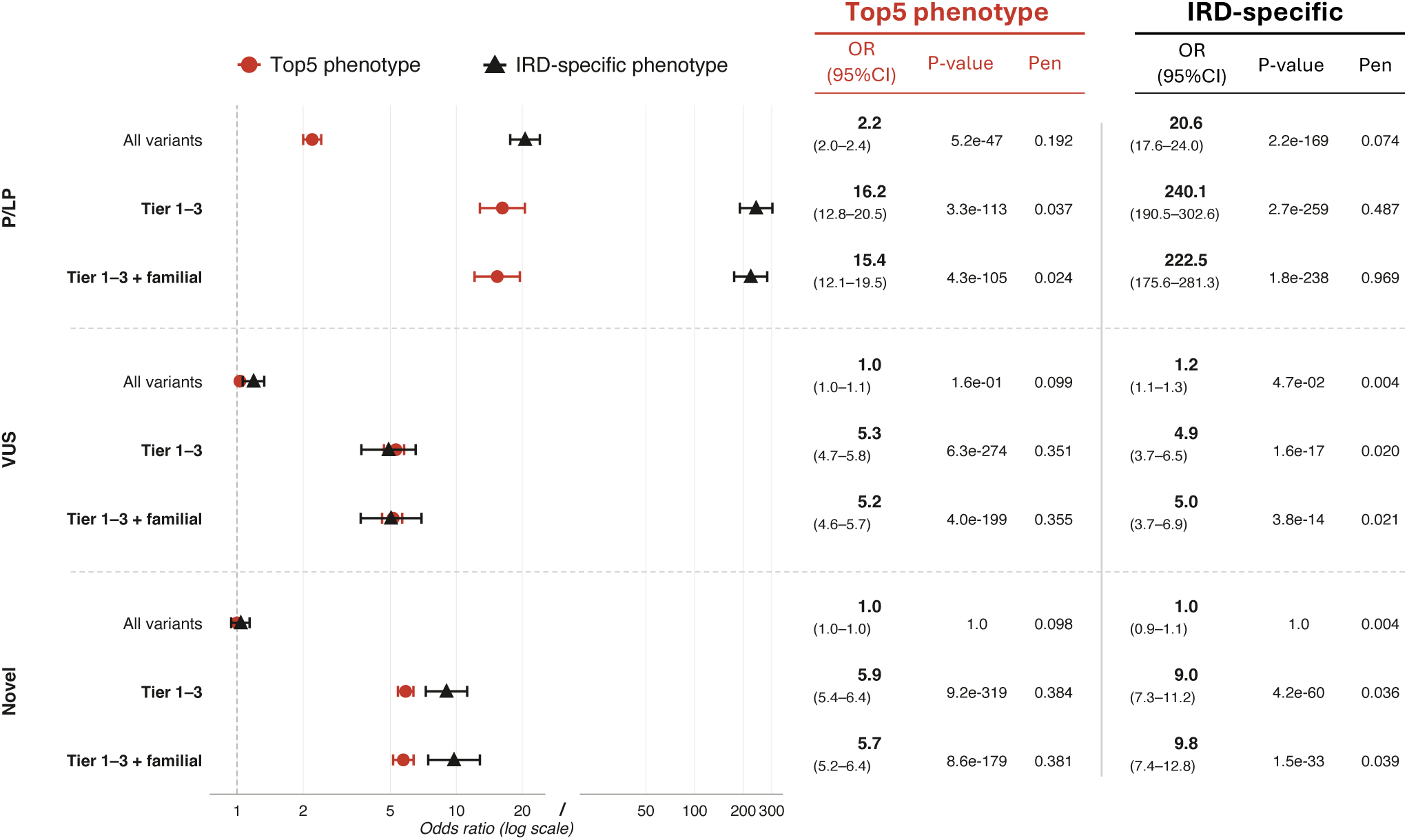
Enrichment of Variant Groups under Broad and IRD-Specific Phenotype Definitions. Odds ratios for enrichment of screened and penetrance-prioritized (Tier 1–3) variants across annotation classes (P/LP (Pathogenic), VUS and novel) are shown using two phenotype definitions: the broader Top5 retinal ICD-10 set and an independent IRD-specific ICD-10 code set curated from a prior population-based study. Enrichment was assessed at the variant-group level using Fisher’s exact test with Bonferroni-adjusted P values.

Importantly, restriction to variants prioritized through our population-based framework (Tiers 1–3) further amplified effect sizes under both phenotype definitions, with ORs increasing from 2.2 to 16.2 for Top5 and from 20.6 to 240.1 for IRD-specific codes. A similar pattern was observed for VUS and novel variants (**Figure 1**). Together, these findings indicate that the Top5-based framework captures IRD-specific disease signal rather than nonspecific ophthalmic burden.

### Familial Structures Support Confidence in IRD Variant Prioritization

The EGP–phenoEGP cohort comprised 75,184 family units and exhibited elevated familial relatedness (mean inbreeding coefficient F = 0.037), substantially higher than that reported in European populations²⁵, including 17,382 families spanning three or more generations. This scale enabled segregation analysis across thousands of independent pedigrees simultaneously — an approach analogous to clinical co-segregation studies but applied at population scale. Integrating the population-based prioritization framework with family structure enabled both validation of prioritized variants through familial concordance and stratification of individuals and families by penetrance tier and observed disease status, distinguishing explained from prevention-relevant genotypes (**Figure 2**). Among the 533 Tier 1–3 prioritized variants, 404 (75.8%) showed familial concordance, defining the population-calibrated IRD panel (**Table 2, Tier 1 variants; Supplementary Appendix, ST8**); of these, 185 were additionally supported by concordance in families with two or more affected individuals. Notably, 91.8% of the 404 panel variants were VUS or novel, underscoring that the majority of variants with the strongest combined population and familial evidence fall outside conventional pathogenic classifications.

**Figure 2.**
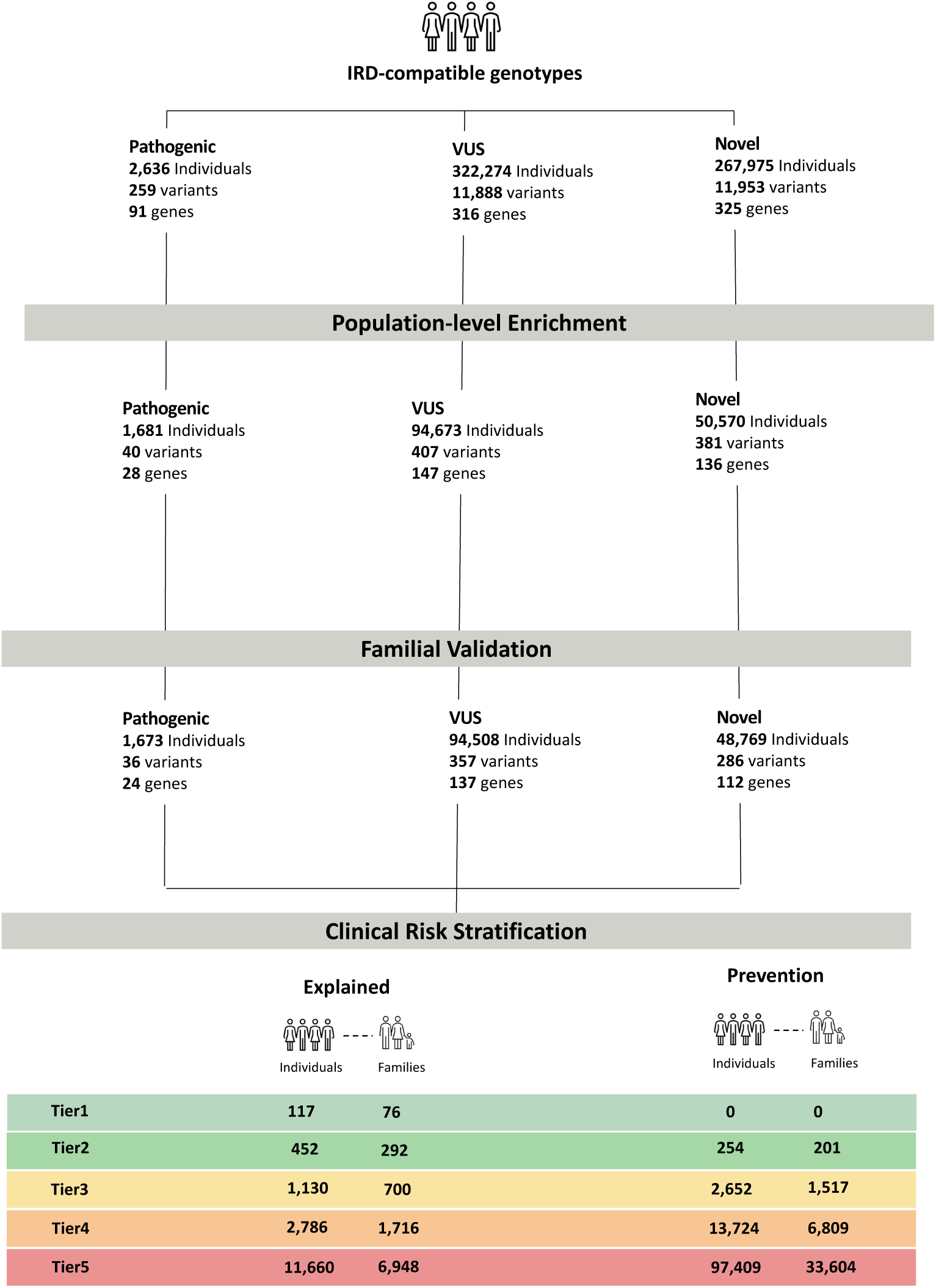
Integrated Population-Based Prioritization and Clinical Stratification of IRD Variants. IRD Variants (P/LP, VUS, Novel) were evaluated through a population-based enrichment framework and subsequent familial validation. Variants showing population enrichment were assigned to penetrance tiers (Tier 1–5) and translated into clinically actionable categories: *Explained* (individuals and families in whom IRD-compatible genotypes are affected with disease) and *Prevention* (IRD-compatible genotypes carriers without documented IRD phenotypes).

**Table 2.** Tier 1 variants in the population-calibrated IRD panel. Variants with complete observed penetrance (100%) identified as part of the population-calibrated IRD panel. Variants are shown in two strata: those validated within reconstructed family units containing two or more affected individuals (upper panel, N=25), and those validated within family units containing at least one affected individual (lower panel, N=23).

| Gene | Variant | Associated disease | Panel | MOI | N | Affected | EGP AF | GnomAD AF |
| --- | --- | --- | --- | --- | --- | --- | --- | --- |
| *Complete penetrance (100%) · Family concordance ≥2 affected individuals · N=25* |  |  |  |  |  |  |  |  |
| **KCNV2 ★** | 9:2718166 G>T | Cone or cone-rod dystrophy | P/LP | AR | 28 | **25** | 8.1×10<sup>-4</sup> | *absent* |
| **ABCA4 ★** | 1:94021339 C>T | Stargardt disease | P/LP | Multiple | 4 | **4** | 2.6×10<sup>-4</sup> | 2.0×10<sup>-5</sup> |
| *ABCA4* | 1:94037335 A>C | Stargardt disease | P/LP | Multiple | 4 | **4** | 3.2×10<sup>-5</sup> | *absent* |
| *INPP5E* | 9:136430391 C>T | Syndromic retinopathy / Joubert syndrome | P/LP | AR | 4 | **4** | 7.9×10<sup>-5</sup> | 6.6×10<sup>-6</sup> |
| *CABP4* | 11:67455483 GC>G | Congenital stationary night blindness | P/LP | AR | 2 | **2** | 1.3×10<sup>-5</sup> | *absent* |
| *MYO7A* | 11:77156990 C>T | Usher syndrome | P/LP | AR | 2 | **2** | 4.0×10<sup>-5</sup> | *absent* |
| *CEP290* | 12:88083863 del | Leber congenital amaurosis | P/LP | AR | 2 | **2** | 5.9×10<sup>-6</sup> | *absent* |
| *CNGB3* | 8:86667131 G>A | Leber congenital amaurosis | P/LP | AR | 2 | **2** | 5.8×10<sup>-5</sup> | 2.0×10<sup>-5</sup> |
| *LRP5* | 11:68386367 C>T | Syndromic retinopathy | VUS | Multiple | 2 | **2** | 2.8×10<sup>-5</sup> | 2.0×10<sup>-5</sup> |
| *PRPF3* | 1:150343315 A>G | Retinitis pigmentosa | VUS | AD | 2 | **2** | 2.4×10<sup>-6</sup> | 3.1×10<sup>-5</sup> |
| **RPE65 Rx** | 1:68431298 T>C | Leber congenital amaurosis | VUS | Multiple | 2 | **2** | 1.5×10<sup>-5</sup> | *absent* |
| *SPP2* | 2:234050985 G>A | Retinitis pigmentosa | VUS | AD | 2 | **2** | 2.4×10<sup>-6</sup> | 6.6×10<sup>-6</sup> |
| *VCAN* | 5:83548072 C>G | Syndromic retinopathy | VUS | AD | 2 | **2** | 2.4×10<sup>-6</sup> | 4.6×10<sup>-5</sup> |
| *RP1L1* | 8:10612496 G>C | Retinitis pigmentosa / macular degeneration | VUS | Multiple | 2 | **2** | 2.7×10<sup>-4</sup> | 3.3×10<sup>-5</sup> |
| *BBS12* | 4:122743596 G>A | Bardet-Biedl syndrome | Novel | AR | 4 | **4** | 5.9×10<sup>-5</sup> | *absent* |
| *MFRP* | 11:119346053 G>A | Other retinopathy (nanophthalmos) | Novel | AR | 3 | **3** | 1.3×10<sup>-4</sup> | 3.3×10<sup>-5</sup> |
| **RPE65 Rx** | 1:68431349 T>G | Leber congenital amaurosis | Novel | Multiple | 3 | **3** | 5.3×10<sup>-4</sup> | 4.6×10<sup>-5</sup> |
| *EFEMP1* | 2:55917877 A>C | Macular degeneration | Novel | AD | 3 | **3** | 3.5×10<sup>-6</sup> | *absent* |
| *COL9A1* | 6:70279562 C>T | Syndromic retinopathy (Stickler syndrome) | Novel | AR | 3 | **3** | 9.9×10<sup>-5</sup> | 1.3×10<sup>-5</sup> |
| *KIF11* | 10:92645374 A>G | Syndromic retinopathy | Novel | AD | 2 | **2** | 2.4×10<sup>-6</sup> | *absent* |
| *DYNC2H1* | 11:103133640 A>G | Other retinopathy | Novel | AR | 2 | **2** | 8.8×10<sup>-5</sup> | 1.3×10<sup>-5</sup> |
| *ABCC6* | 16:16161472 C>T | Syndromic retinopathy (pseudoxanthoma elasticum) | Novel | Multiple | 2 | **2** | 1.4×10<sup>-4</sup> | *absent* |
| *DRAM2* | 1:111139519 A>G | Macular degeneration | Novel | AR | 2 | **2** | 7.9×10<sup>-5</sup> | 2.0×10<sup>-5</sup> |
| *PRDM13* | 6:99613249 C>A | Chorioretinal atrophy or degeneration | Novel | AD | 2 | **2** | 2.4×10<sup>-6</sup> | *absent* |
| *MT-ATP6* | m.8767 A>G | Mitochondrial retinopathy | Novel | MT | 2 | **2** | 1.06E-05 | *absent* |
| **Complete penetrance (100%) · Family concordance ≥1 affected individual · N=23** |  |  |  |  |  |  |  |  |
| *CRB1* | 1:197442248 T>G | Leber congenital amaurosis / retinitis pigmentosa | P/LP | Multiple | 5 | 5 | 2.4×10<sup>-4</sup> | absent |
| *ABCA4* | 1:94025021 G>A | Stargardt disease | P/LP | Multiple | 2 | 2 | 3.4×10<sup>-5</sup> | absent |
| *PDE6B* | 4:653950 C>A | Retinitis pigmentosa | P/LP | Multiple | 2 | 2 | 9.7×10<sup>-5</sup> | 1.3×10<sup>-5</sup> |
| *RPGR* | X:38286592 del | Retinitis pigmentosa / cone-rod dystrophy | P/LP | XL | 2 | 2 | 1.06E-05 | absent |
| *CABP4* | 11:67452532 G>A | Congenital stationary night blindness | VUS | AR | 2 | 2 | 7.0×10<sup>-4</sup> | 2.4×10<sup>-4</sup> |
| *GNPTG* | 16:1362047 C>G | Syndromic retinopathy (mucopolipidosis) | VUS | AR | 2 | 2 | 6.3×10<sup>-5</sup> | 6.6×10<sup>-6</sup> |
| *CNGB1* | 16:57887986 C>T | Retinitis pigmentosa | VUS | AR | 2 | 2 | 4.2×10<sup>-4</sup> | 2.0×10<sup>-4</sup> |
| *MFN2* | 1:11996289 G>A | Optic atrophy / syndromic retinopathy | VUS | AD | 2 | 2 | 2.4×10<sup>-6</sup> | 2.6×10<sup>-5</sup> |
| *FLVCR1* | 1:212888562 A>T | Syndromic retinopathy | VUS | AR | 2 | 2 | 2.3×10<sup>-4</sup> | 5.9×10<sup>-5</sup> |
| *ABHD12* | 20:25308042 G>A | Usher syndrome / PHARC syndrome | VUS | AR | 2 | 2 | 1.0×10<sup>-4</sup> | 1.3×10<sup>-5</sup> |
| *CFAP410* | 21:44331843 G>A | Cone or cone-rod dystrophy | VUS | AR | 2 | 2 | 1.4×10<sup>-4</sup> | 3.6×10<sup>-4</sup> |
| *GUCA1B* | 6:42194765 G>A | Retinitis pigmentosa / macular degeneration | VUS | AD | 2 | 2 | 2.4×10<sup>-6</sup> | 1.3×10<sup>-5</sup> |
| *IMPG1* | 6:76005322 T>C | Retinitis pigmentosa / macular degeneration | VUS | Multiple | 2 | 2 | 1.1×10<sup>-4</sup> | 1.3×10<sup>-4</sup> |
| *TPPA* | 8:63059767 del | Syndromic retinopathy (ataxia with vit E deficiency) | VUS | AR | 2 | 2 | 3.7×10<sup>-4</sup> | 1.3×10<sup>-4</sup> |
| *GDF6* | 8:96145387 C>T | Leber congenital amaurosis | VUS | AR | 2 | 2 | 2.9×10<sup>-4</sup> | absent |
| *TULP1* | 6:35498367 A>G | Leber congenital amaurosis / retinitis pigmentosa | Novel | AR | 4 | 4 | 1.0×10<sup>-4</sup> | absent |
| *VWA8* | 13:41721477 G>C | Retinitis pigmentosa | Novel | AD | 3 | 3 | 3.5×10<sup>-6</sup> | absent |
| *TRPM1* | 15:31068055 del | Congenital stationary night blindness | Novel | AR | 3 | 3 | 2.2×10<sup>-4</sup> | absent |
| *EFEMP1* | 2:55881621 G>T | Macular degeneration | Novel | AD | 2 | 2 | 2.4×10<sup>-6</sup> | absent |
| *GRM6* | 5:178994469 A>C | Congenital stationary night blindness | Novel | AR | 2 | 2 | 1.8×10<sup>-4</sup> | absent |
| *RIMS2* | 8:104015442 T>C | Syndromic retinopathy | Novel | AR | 2 | 2 | 7.9×10<sup>-5</sup> | 5.9×10<sup>-5</sup> |
| *CEP78* | 9:78236592 TG>T | Cone or cone-rod dystrophy | Novel | AR | 2 | 2 | 1.1×10<sup>-5</sup> | absent |
| *RPGR* | X:38286110 T>G | Retinitis pigmentosa / cone-rod dystrophy | Novel | XL | 2 | 2 | 1.06E-05 | absent |

★ Top overall enrichment signal across all 24,100 screened variants. Rx: RPE65 variants — gene therapy eligible (voretigene neparvovec). MOI: mode of inheritance; AR: autosomal recessive; AD: autosomal dominant; XL: X-linked; Mito: mitochondrial. EGP AF: allele frequency in the Emirati Genome Program. gnomAD AF: overall allele frequency in gnomAD genome sequences; 'absent' = not observed.

Segregation within reconstructed pedigrees was consistent with mode of inheritance patterns across novel variants in *RPE65*, *BBS12*, *PRDM13*, *COL9A1*, *MFRP*, and *EFEMP1* (**Supplementary Appendix, SF5, SF6**), including a VUS in *RPE65* segregating with autosomal recessive inheritance and a novel variant in *BBS12* subsequently annotated as pathogenic in ClinVar, underscoring the value of our population-calibrated prioritization. However, these pedigrees also revealed interpretive challenges inherent to EHR-derived phenotypes: in the *RPE65* family, a Tier 1 novel variant (chr1:68431349:T>G; penetrance 100%, 3/3 affected) showed complete concordance among individuals with IRD-compatible genotypes, yet an older heterozygous carrier also carried a retinal diagnosis — possibly reflecting dominant effects, age-related retinal disease, or a distinct etiology captured under the broad Top5 phenotype definition. In the *ACBD5* family, a Tier 2 novel variant (chr10:27199037:A>C; penetrance 62.5%, 5/8 affected) showed an unaffected individual with an IRD-compatible genotype alongside multiple affected offspring. Both patterns underscore the need for dedicated clinical phenotyping to confirm variant pathogenicity beyond population-based prioritization.

### From Clinical Observation to Population-Level Risk

Our population-based variant prioritization framework identified two co-occurring ABCA4 variants — c.5882G>A and c.2570T>C — among the prioritized variants in the panel (**Supplementary Appendix, ST4**). When evaluated independently, c.5882G>A appeared to have substantially higher penetrance (∼80%, Tier 2) than c.2570T>C (∼20%, Tier 4), which might suggest that the former drives disease risk as a single variant. However, population-scale genotyping revealed that every individual homozygous for c.5882G>A was also homozygous for c.2570T>C, indicating that the apparent signal of c.5882G>A reflects a shared haplotype rather than an independent pathogenic effect (**Figure 3A**). This recurrent haplotype had been previously recognized through clinical evaluation of affected Emirati families with Stargardt disease^26^; our population-scale framework not only confirms that the association is genuine and free of ascertainment bias, but resolves its underlying haplotypic architecture, demonstrating that neither variant can be interpreted in isolation.

**Figure 3.**
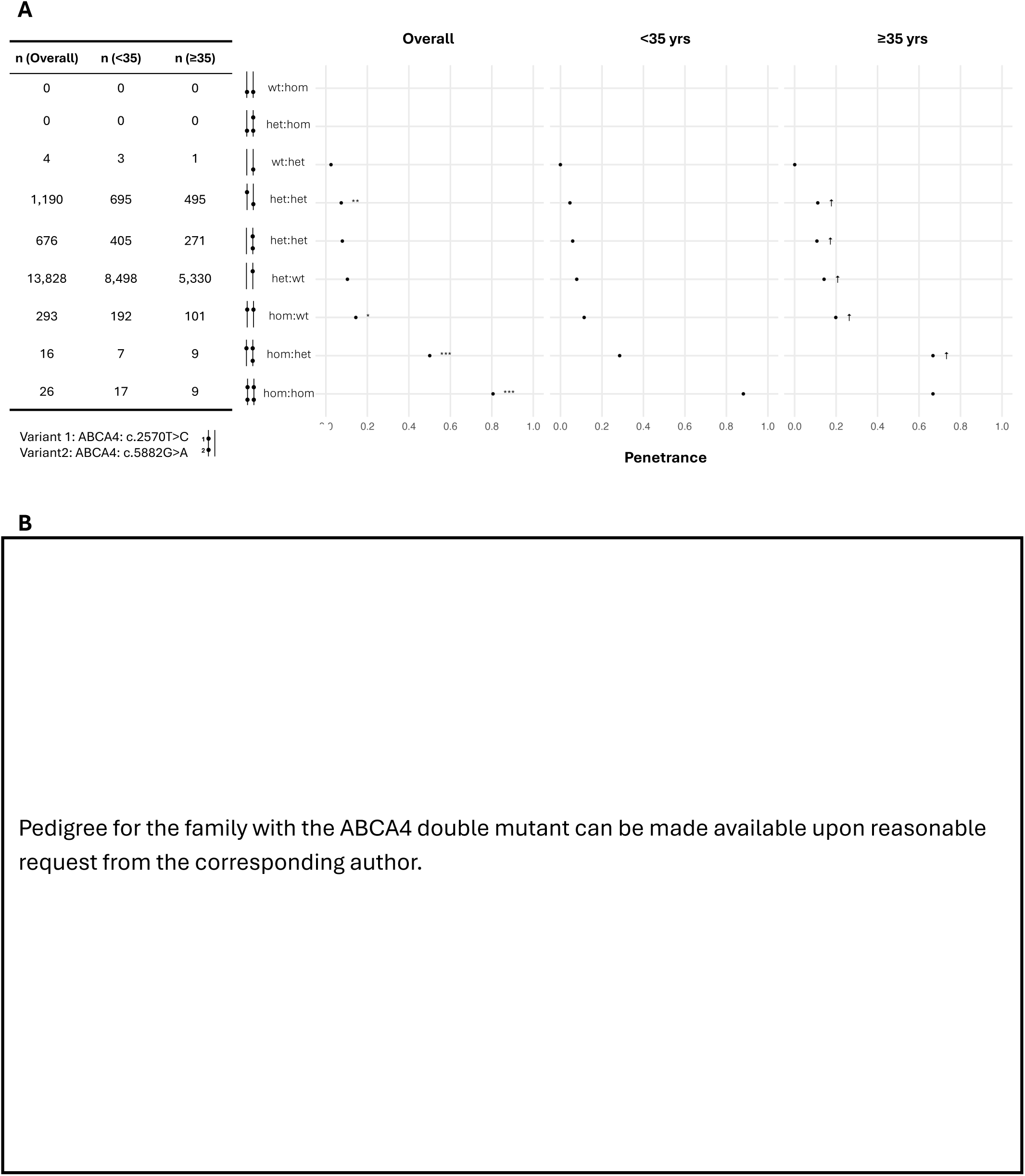
Haplotype-Level Analysis of a Recurrent ABCA4 Locus. Panel. **A** shows population-level penetrance estimates for two *ABCA4* variants evaluated independently, and in combination as a shared haplotype, stratified by age group. **Panel B** shows segregation of the ABCA4 haplotype within an extended family network, illustrating concordance between double-homozygosity and IRD phenotypes.

Family-based analysis identified an extended family network harboring this haplotype, with three double-homozygous individuals, all of whom were affected (**Figure 3B**). Double-homozygous carriers showed significantly increased runs of homozygosity spanning the *ABCA4* locus compared with random genomic regions, whereas no such enrichment was observed among cis-heterozygous carriers or noncarriers, consistent with localized autozygosity underlying the haplotype (**Supplementary Appendix, SF7 and ST9**).

## Discussion

In this population-scale study, we integrated WGS, longitudinal EHR, and reconstructed multigenerational family structures to develop a scalable framework for variant interpretation in IRD. Several key findings emerged. Penetrance was highly heterogeneous across IRD-associated variants; a subset of variants classified as P/LP lacked population-level support; reproducible disease risk was observed among subsets of variants currently classified as uncertain or novel; and incorporation of family structure strengthened inference and enabled validation of high-risk variants. Collectively, these results demonstrate that variant pathogenicity cannot be assumed constant across populations and that population-calibrated interpretation is a necessary complement to assertion-based classification when genomic screening is deployed at national scale.

Large biobank analyses have challenged the assumption that Mendelian disorders exhibit near-complete penetrance^4,8^. In a recent population-scale study of IRD-compatible genotypes, penetrance ranged from approximately 9% to 28%, substantially lower than historical expectations^6^. Our findings confirm this observation and further demonstrate heterogeneity at the variant level. Similar patterns of reduced penetrance have been reported across other Mendelian conditions in population-based cohorts^8,27,28^, suggesting that variable clinical expression is a generalizable property of rare disease variants. We extend prior work by moving beyond penetrance estimation toward operational recalibration, integrating enrichment testing, penetrance tiering, and familial concordance within a unified framework that distinguishes unsupported annotations from reproducible population-level risk alleles.

Important implications arise for variants annotated as P/LP. Although these variants demonstrated the highest proportional enrichment overall, a subset lacked population-level support when sufficient carrier representation was available. Among P/LP variants observed in at least 10 individuals with IRD-compatible genotypes, approximately 10% failed to show significant enrichment, and 3% exhibited null penetrance despite adequate carrier counts. These findings underscore that pathogenicity annotations derived from clinically ascertained cohorts may not uniformly translate to general populations^29^. In population screening context, classification alone may overestimate risk without calibration against real-world disease expression.

Conversely, among more than 23,000 VUS and novel variants evaluated, only a focused subset met criteria for significant enrichment and moderate-to-high penetrance. This reduction from thousands of uncertain candidates to a concentrated group of high-risk alleles illustrates the framework’s ability to extract clinically meaningful signal from background variation. For VUS, such evidence may inform reclassification; for novel variants, it enables systematic identification of previously unclassified risk alleles with population-level disease support, particularly in historically understudied populations. This bidirectional recalibration —downgrading unsupported P/LP variants while elevating high-risk uncertain and novel alleles — has direct clinical consequences. Among the strongest population-level signals were P/LP variants in *KCNV2* and *ABCA4*, both absent from gnomAD genome sequences and enriched more than 100-fold in the Emirati cohort, providing actionable evidence to guide clinical prioritization and ensure that the most population-relevant variants are reflected in regional diagnostic panels. The framework also identified novel variants subsequently validated through independent evidence, including a *BBS12* variant later reclassified as pathogenic in ClinVar, illustrating the potential for population-calibrated prioritization to anticipate clinical classification. The stakes of misclassification are perhaps most sharply illustrated by *RPE65*: our framework identified both a VUS and a novel variant with complete penetrance and family-based concordance across multiple affected individuals. *RPE65*-associated IRD is among the forms of IRDs with a regulatory-approved gene therapy^30^— voretigene neparvovec — where accurate variant classification directly determines treatment eligibility. Population-scale evidence of complete penetrance, confirmed across independent pedigrees and absent from global reference databases, provides direct grounds for reclassification and clinical re-evaluation of these individuals.

Unlike prior population-scale analyses that evaluated penetrance primarily at the individual level, our framework incorporates reconstructed family units to approximate key elements of clinical genetic reasoning at scale, an approach analogous to clinical co-segregation studies but applied simultaneously across thousands of independent pedigrees. Although formal segregation analyses were not performed, evaluation of concordance within reconstructed families provided an additional layer of evidence, distinguishing variants that recur consistently among affected relatives from those enriched only in aggregate population data.

Importantly, population-scale observations of penetrance do not imply that clinically observed associations weaken in unselected cohorts. In certain contexts, strong clinical signals are reinforced when evaluated at scale. The *ABCA4* haplotype illustrates this relationship directly. First recognized through clinical evaluation of affected Emirati families with Stargardt disease^26^, this recurrent haplotype demonstrated substantial disease association within the population-scale dataset. Interpretation within its shared haplotypic configuration revealed higher penetrance, and concordant occurrence within reconstructed family units further strengthened inference, reinforcing pathogenic relevance.

This study has limitations inherent to population-scale analyses. Phenotype ascertainment relied on routinely collected EHR data and ICD-10 coding, which may incompletely capture disease onset or severity. The use of a broad retinal phenotype set, although reflecting clinical heterogeneity, may capture nonspecific visual conditions; family-level pedigree patterns in which non-carriers carry retinal diagnoses, or in which at-risk parents remain unaffected, highlight that EHR-derived phenotypes require dedicated clinical phenotyping to confirm or refute variant pathogenicity in specific families. Enrichment analyses may remain susceptible to residual false-positive findings; however, prioritization required concordant evidence across enrichment testing, penetrance tiering, and familial concordance, mitigating spurious associations. Although the analytic framework is broadly applicable, replication in other populations will be necessary to assess generalizability of specific variant effects.

These findings have direct implications for population genomic screening and the clinical interpretation of genetic testing for rare diseases. Variant panels constructed solely from assertion-based classifications risk both overestimating risk for variants lacking population-level support and failing to identify high-penetrance alleles embedded within uncertain or previously unreported categories. The framework described here, integrating enrichment testing, penetrance tiering, and familial concordance against longitudinal EHR data, offers a practical model for population-calibrated panel construction applicable wherever population-scale genomic and health record data are available. The identification of *RPE65* carriers with complete-penetrance variants — individuals who would not qualify for gene therapy referral under current classification frameworks — illustrates what is at stake: not a refinement of existing practice, but access to treatment. As genome sequencing expands into diverse and previously uncharacterised populations, the gap between assertion-based classification and real-world disease expression will widen. Closing that gap requires population-scale calibration to be treated not as a refinement of existing frameworks but as a foundational step in responsible genomic screening.

## Supplementary Materials and Methods

### Study Population

Participants were enrolled in the EGP; a nationwide initiative launched in 2019 by the Department of Health of Abu Dhabi (DOH) to sequence the genomes of Emirati citizens for research and precision medicine. Our current analysis included WGS that had been completed for 504,000 individuals recruited between January 2020 and October 2024. Additional details on sequencing platforms and variant curation are provided in the following section.

Among these, 426,382 individuals had linked phenoEGP available through phenoEGP (EGP-phenoEGP cohort). Individuals without phenoEGP data or missing essential demographic information were excluded from phenotype-based analyses.

### WGS, Alignment and Variant Calling

WGS was performed to a target depth of 30x in one of three platforms: Illumina, MGI, or Oxford Nanopore Technologies (ONT). Sequencing reads were aligned to the human reference genome assembly GRCh38 using platform-specific pipelines: the DRAGEN Bio-IT Platform 3.9 (Illumina)^31^, BWA-MEM algorithm (MGI)^32^ and Sentieon’s acceleration of Minimap2 (ONT)^33^.

Variant calling was conducted using DRAGEN Bio-IT Platform 3.9 for Illumina data^31^, Sentieon’s accelerated implementation of GATK HaplotypeCaller for MGI data^34^, and Clair2 for ONT data^35^. Variant calls from all platforms were normalized to a unified representation and merged into a harmonized multi-platform variant dataset for downstream analysis.

### Sample– and Variant-Level Quality Control

Sample-level quality control was applied to exclude individuals with low sequencing coverage (<26x), sex discordance, low mapping rate (<70%), or signs of contamination (contamination >1% or Het/Hom ratio ≥2.5). Variant-level QC excluded variants with low genotype quality or depth, using platform-specific thresholds as outlined in (**Supplementary Appendix, ST10**). Only high-confidence variants and samples passing all filters were retained for downstream analyses.

As part of the genomic screening workflow, additional cross-platform enrichment analyses computed using Fisher’s exact tests were performed to detect variants disproportionately over– or under-called in specific platforms. These were used to exclude variants likely affected by platform-specific technical bias from downstream analyses.

### Kinship Inference and Families Construction

We first generated a genome-wide dataset in PLINK format simulating Illumina Global Screening Array-24 v3.0 (GSA)^20^, which includes 608,896 variants. This array-based dataset was constructed for 504,000 individuals in the EGP and served as input for kinship analyses. To assess levels of autozygosity and consanguinity, we computed individual-level inbreeding coefficients (F-statistics) using PLINK, version 1.9^36^, with the ––het flag.

We defined a family as a set of individuals connected through inferred biological relationships based on genome-wide identity-by-descent (IBD) estimates. Kinship inference was performed using KING, version 2.2.7^21^, which calculates pairwise kinship coefficients and classifies relationship types based on autosomal genotype data. Relationship categories included: Parent-offspring (PO): kinship coefficient ≈ 0.25 and IBD2 ≈ 0, Full siblings (FS): kinship coefficient ≈ 0.25 and IBD2 > 0, Second-degree relatives (e.g., grandparent–grandchild, avuncular, half-siblings): kinship coefficient ≈ 0.125, Third-degree relatives: kinship coefficient ≈ 0.0625.

To identify multigenerational family structures, we focused exclusively on PO relationships inferred using KING. Pairwise PO links were converted into a graph using the igraph package in R^37^, where nodes represent individuals and edges represent directed PO relationships. Family units were defined as multigenerational clusters anchored by the earliest inferred generation (G1) and including direct descendants (G2 and G3), enabling inheritance-aware variant interpretation and family-based enrichment analyses. PO pairs were used to assign pedigree directionality using available age and sex information, assuming the older individual to be the parent. Pedigrees were constructed using the kinship2 package in R^38^, supporting visualization.

### Variant Annotation and Functional Consequence Prediction

To identify potentially novel IRD-associated variants we extracted all polymorphic sites located within RetNet genes (339 genes; accessed April 2025) across the EGP-phenoEGP cohort. Variant MAF was calculated from observed allele counts. Variants with MAF <1% were carried forward for functional annotation. These variants were annotated using the Ensembl VEP^23^, version 112.0, with reference genome GRCh38. Predicted functional consequences were assigned based on Ensembl gene models and transcript annotations.

For downstream analyses, we retained variants annotated as having moderate or high predicted functional impact, including missense, nonsense, frameshift, and canonical splice-site variants, and absent from ClinVar. These variants were designated as novel and evaluated as previously unclassified candidates for IRD association.

### Electronic Health Record Processing and PheWAS

EHRs were accessed through Malaffi (Arabic for “my file”), Abu Dhabi’s health information exchange (HIE) operated by Abu Dhabi Health Data Services (ADHDS), an M42 company established in partnership with the Department of Health Abu Dhabi (DOH). Malaffi provides centralized, longitudinal clinical records for patients across public and private healthcare facilities in the Emirate. For this study, we used the subset of Malaffi data specifically linked to participants in the EGP hereafter referred to as phenoEGP. PhenoEGP comprises 10 years of longitudinal clinical data (2013–2024), including diagnoses, procedures, prescriptions, and laboratory results, integrated with genomic data from the EGP to enable large-scale genotype–phenotype analyses.

Phenotypic associations were evaluated using a PheWAS based on ICD-10 diagnoses derived from phenoEGP. Diagnoses were encoded using ICD-10 and restricted to codes within Chapter VII (Diseases of the Eye and Adnexa; H00–H59). To enrich for rare, potentially Mendelian retinal phenotypes and reduce noise from common, nonspecific diagnoses, ICD-10 codes with a population prevalence ≥1% in the EGP–phenoEGP cohort were excluded (n= 50). This filtering resulted in a refined set of 2,323 ICD-10 codes used for downstream association analyses. For each variant, ICD-10 associations were tested using Fisher’s exact test, comparing variant carriers with non-carriers, applying a continuity correction and estimating odds ratios, risk differences, and P values. Multiple testing correction was applied using the Benjamini–Hochberg false discovery rate (FDR) procedure within each analysis.

To identify retinal phenotype groupings enriched among IRD variant carriers, association signals were aggregated across variants using a weighted enrichment framework incorporating effect size and statistical significance. ICD-10 subchapters were ranked by cumulative enrichment, and the five most consistently enriched subchapters, H35 (retinal disorders), H54 (visual impairment), H55 (nystagmus), H50 (strabismus), and H53 (visual disturbances), were selected for downstream analyses. Together, these subchapters comprised 359 ICD-10 codes and were collectively referred to as the Top5 phenotypes, which served as proxies for IRD manifestations in all population-level enrichment and penetrance analyses.

### Definition of IRD-compatible genotypes

Variants were extracted from whole-genome variant call files using bcftools (v1.21). Carrier status was determined for each individual (heterozygous: one alternate allele; homozygous: two alternate alleles). Individuals were classified as carrying IRD-compatible genotypes based on variant zygosity and gene-level mode of inheritance. IRD-compatible genotypes were defined as homozygous carriers of a variant, regardless of reported mode of inheritance, or heterozygous carriers of variants in genes with autosomal dominant inheritance.

To account for uncertainty in inheritance annotation, possible IRD-compatible genotypes were defined as heterozygous carriers of variants in genes with multiple or unknown modes of inheritance. These genotypes were included in sensitivity analyses to evaluate whether such variants exhibited penetrance patterns consistent with dominant effects at the population level.

### Population-Based Enrichment and Penetrance Estimation

Population-based enrichment was assessed at the variant level using Fisher’s exact test, comparing the proportion of individuals with IRD phenotypes among genotype-compatible variant carriers to non-carriers within the EGP–phenoEGP cohort. IRD cases were defined using the Top5 retinal phenotype set, as described above.

For each variant, carrier risk and non-carrier risk were calculated as the proportion of individuals with IRD phenotypes within each group, and the risk difference (RD) was defined as the difference between these proportions. Variants with RD > 0 and nominal evidence of enrichment (P < 0.05) were carried forward for penetrance estimation. No multiple-testing correction was applied at this stage, as enrichment was used as a directional screening criterion rather than a final inference test.

Penetrance was calculated as the proportion of genotype-compatible carriers with at least one recorded Top5 retinal phenotype. Variants were assigned to penetrance tiers based on observed penetrance thresholds (Tier 1–5). Prioritized variants were defined as those with an observed penetrance of ≥25% (Tier1-3). Variants observed in at least 10 individuals with IRD-compatible genotypes but with no IRD cases were designated as having null penetrance and were evaluated separately from penetrance-tiered variants.

To identify variants with the strongest combined enrichment signal, a composite score was derived for each variant observed in IRD-compatible genotypes as the product of the carrier risk difference and −log₁₀(P value), and variants were ranked accordingly.

### Age-Stratified Analyses

To account for the progressive onset of many IRDs, we performed an age-stratified analysis restricted to individuals aged ≥35 years. This threshold was selected to increase sensitivity for detecting variants associated with later-onset or slowly progressive retinal phenotypes. Within the ≥35-year subset, we re-applied the population-based variant prioritization framework, including variant-level enrichment testing, penetrance estimation among IRD-compatible genotype carriers, and assignment to penetrance tiers. All definitions, thresholds, and criteria were identical to those used in the primary analysis. Variants meeting Tier 1–3 penetrance criteria in the ≥35-year subset but not in the full cohort were identified as age-dependent prioritized variants.

### Robustness Analysis Using IRD-Specific ICD-10 Codes

To assess whether variants prioritized using the broader Top5 retinal phenotype captured disease signal under a stricter clinical definition, we performed an independent enrichment analysis using an IRD-specific set of 35 ICD-10 codes curated from a recent population-based study^6^ (referred to as the “IRD set” in the original publication).

Enrichment was evaluated at the level of variant groups rather than individual variants. Specifically, we assessed enrichment among IRD cases for: (i) all screened variants stratified by annotation class (P/LP, VUS, and novel), and (ii) penetrance-prioritized variants (Tiers 1–3), stratified by the same annotation classes. For each variant group, enrichment relative to non-carriers was assessed using Fisher’s exact test with continuity correction, and statistical significance was determined using Bonferroni-adjusted P values.

The IRD-specific ICD-10 codes were not used in enrichment testing or penetrance tier assignment, which were based on the broader Top5 retinal phenotype definition. This analysis therefore evaluated enrichment under an independently curated, more specific phenotype definition.

### Family concordance analyses

To evaluate whether population-prioritized variants were supported by familial disease patterns, we performed a family-based concordance analysis across reconstructed family units.

For each prioritized variant (Tiers 1–3), we identified family units containing at least one IRD case who carried the variant and excluded family–variant pairs in which one or more IRD cases were non-carriers. Among the remaining family units, when multiple prioritized variants were present, the variant with the strongest population-level support was designated as the best-explaining variant.

Population-level support was defined hierarchically, ranking variants first by penetrance tier (Tier 1 > Tier 2 > Tier 3 > Tier 4 > Tier 5) and, when tiers were identical, by variant class (pathogenic/likely pathogenic > variants of uncertain significance > novel variants). A variant was considered to show familial concordance if it was designated as the best-explaining variant in at least one family unit containing IRD cases.

### Runs of Homozygosity Analysis at the ABCA4 Locus

Genotype phasing was performed using Beagle (version 22Jul22.46e) to infer haplotype structure across the ABCA4 locus Phased genotypes were used to determine whether the two ABCA4 variants occurred in cis (on the same chromosome) or trans (on opposite chromosomes), a distinction critical for evaluating biallelic pathogenicity and for downstream runs-of-homozygosity analyses.

To evaluate whether the recurrent ABCA4 haplotype was associated with localized autozygosity, we performed a runs of homozygosity (ROH) analysis focused on the ABCA4 genomic region on chromosome 1 (chr1:93.9–94.2 Mb, GRCh38). ROH segments were identified using PLINK (version 1.9) with standard parameters optimized for detecting long autozygous segments (--homozyg, minimum 50 SNPs, minimum length 1.5 Mb, maximum gap 1 Mb). ROH calling was restricted to individuals included in the ABCA4 haplotype analysis.

Individuals were grouped into three categories based on ABCA4 genotype: (1) double-homozygous carriers of the ABCA4 haplotype, (2) cis-heterozygous carriers of the haplotype, and (3) unrelated noncarrier controls. Control individuals were randomly sampled from the EGP-phenoEGP cohort and filtered to exclude related individuals based on KING-derived kinship estimates, ensuring independence from carrier families.

For each individual, the proportion of the ABCA4 locus covered by ROH segments was calculated. To assess whether ROH enrichment at the ABCA4 locus exceeded background levels, we generated 500 matched-length control windows randomly sampled across chromosome 1, excluding a 2-Mb buffer around the ABCA4 region. ROH coverage was calculated for each control window using the same approach.

Group-level comparisons were performed by contrasting mean ROH coverage at the ABCA4 locus against the empirical distribution derived from the random control windows. Empirical P values were computed as the proportion of control windows with equal or greater ROH coverage than observed at the ABCA4 locus. Visualization of ROH segments across chromosome 1 was performed using the R package karyoploteR, enabling locus-centered representation of individual-level autozygous segments.

## Data Availability

All data used in this study are part of the Emirati Genome Program (EGP) and phenoEGP, governed by the Department of Health Abu Dhabi. Access may be granted upon reasonable request and institutional approval.

