## Supplementary Figures for "Pathogenicity Reassessment and Novel Variant Discovery in Inherited Retinal Disease through Population-Scale Genomics in the United Arab Emirates" (from another version of this preprint: version 1, 2025-10-19)

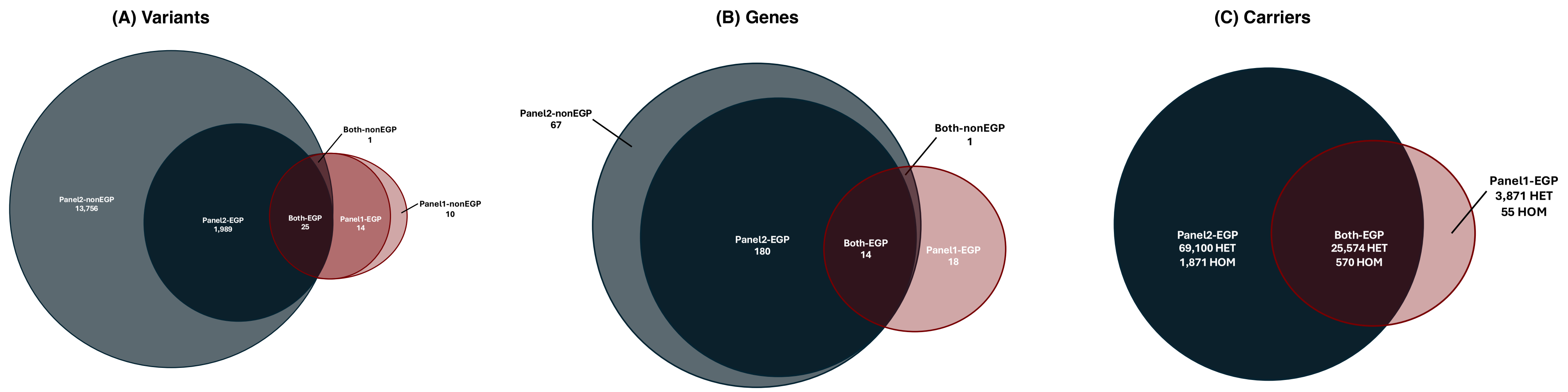


**Supplementary Figure 1. Overlap between Panel I and Panel II in terms of screened variants, genes, and identified carriers within the EGP cohort.**

(A) Variants: Of the 15,795 variants screened across both panels, 2,028 were observed among carriers in the EGP cohort. (B) Genes: The screened variants spanned 251 genes in total, with 204 genes observed in carriers within the EGP. (C) Carriers: Across 504,000 individuals, we identified 92,476 heterozygotes (18.3%) and 2,460 homozygotes (0.49%).

Restricted

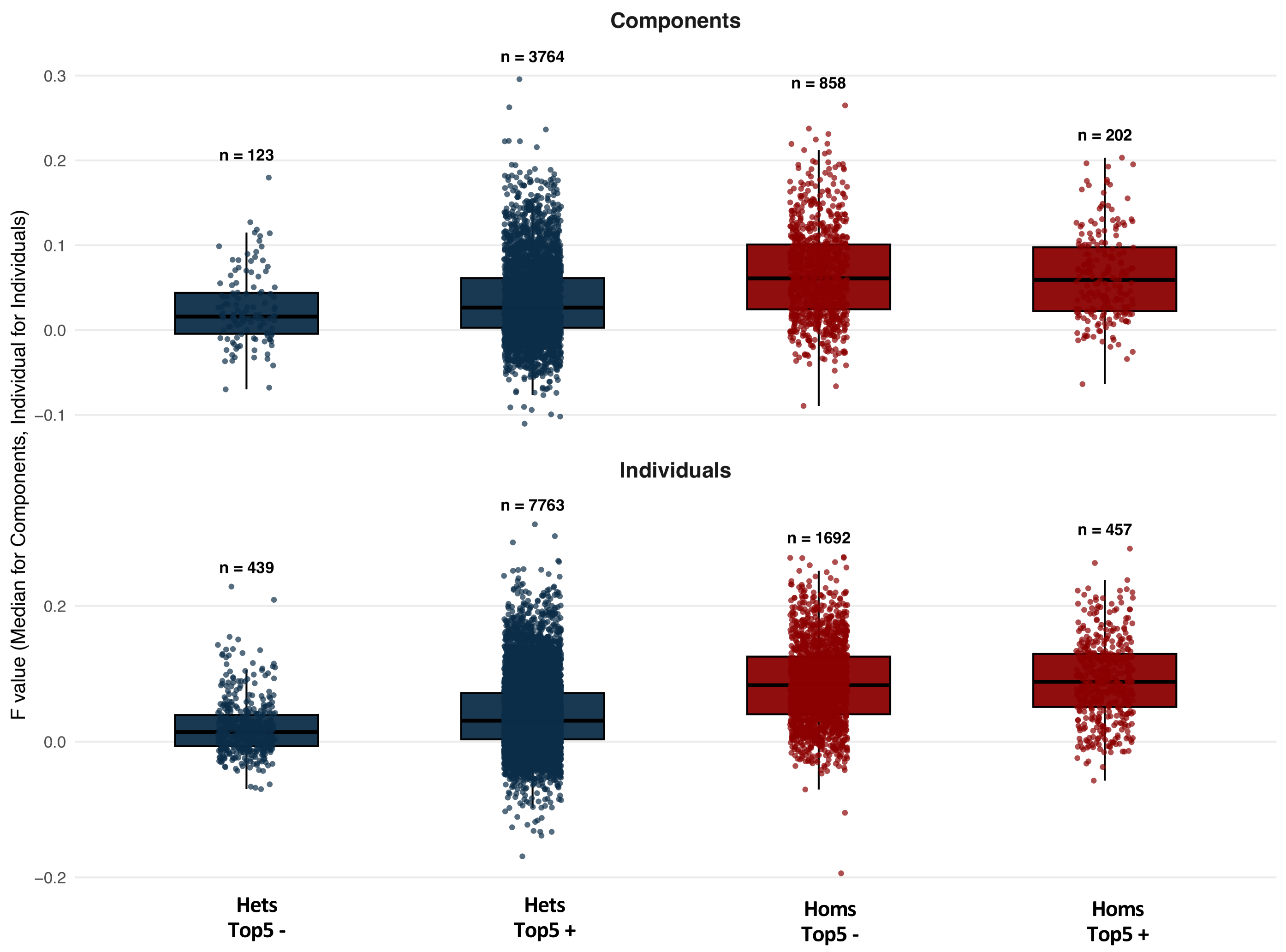
Components
n = 123
n = 3764
n = 858
n = 202
0.3
0.2
0.1
0.0
−0.1
Individuals
n = 439
n = 7763
n = 1692
n = 457
0.2
0.0
−0.2
F value (Median for Components, Individual for Individuals)
Hets Top5 -
Hets Top5 +
Homs Top5 -
Homs Top5 +

**Supplementary Figure 2. Inbreeding Coefficients (F values) of Carrier Groups at the Component and Individual Levels.**

Boxplots show the distribution of inbreeding coefficients (F values, estimated with plink --het) across four carrier groups, stratified by analysis type: Components (top, median F per genetically defined family component) and Individuals (bottom, per-individual F). Carrier groups were defined as: (1) Only heterozygotes without top five phenotypes (dark blue), (2) Only heterozygotes with top five phenotypes (light blue), (3) Homozygotes without top five phenotypes (dark red), and (4) Homozygotes with top five phenotypes (light red). The top five phenotypes correspond to the most frequent IRD-related ICD-10 subchapters (H35, H54, H55, H50, H53). For clarity, heterozygous carriers of autosomal recessive (AR) genes were excluded, as penetrance estimates based on single heterozygotes are not informative for recessive inheritance. Points represent individual components (top) or individuals (bottom), with sample sizes (n) indicated above each distribution.

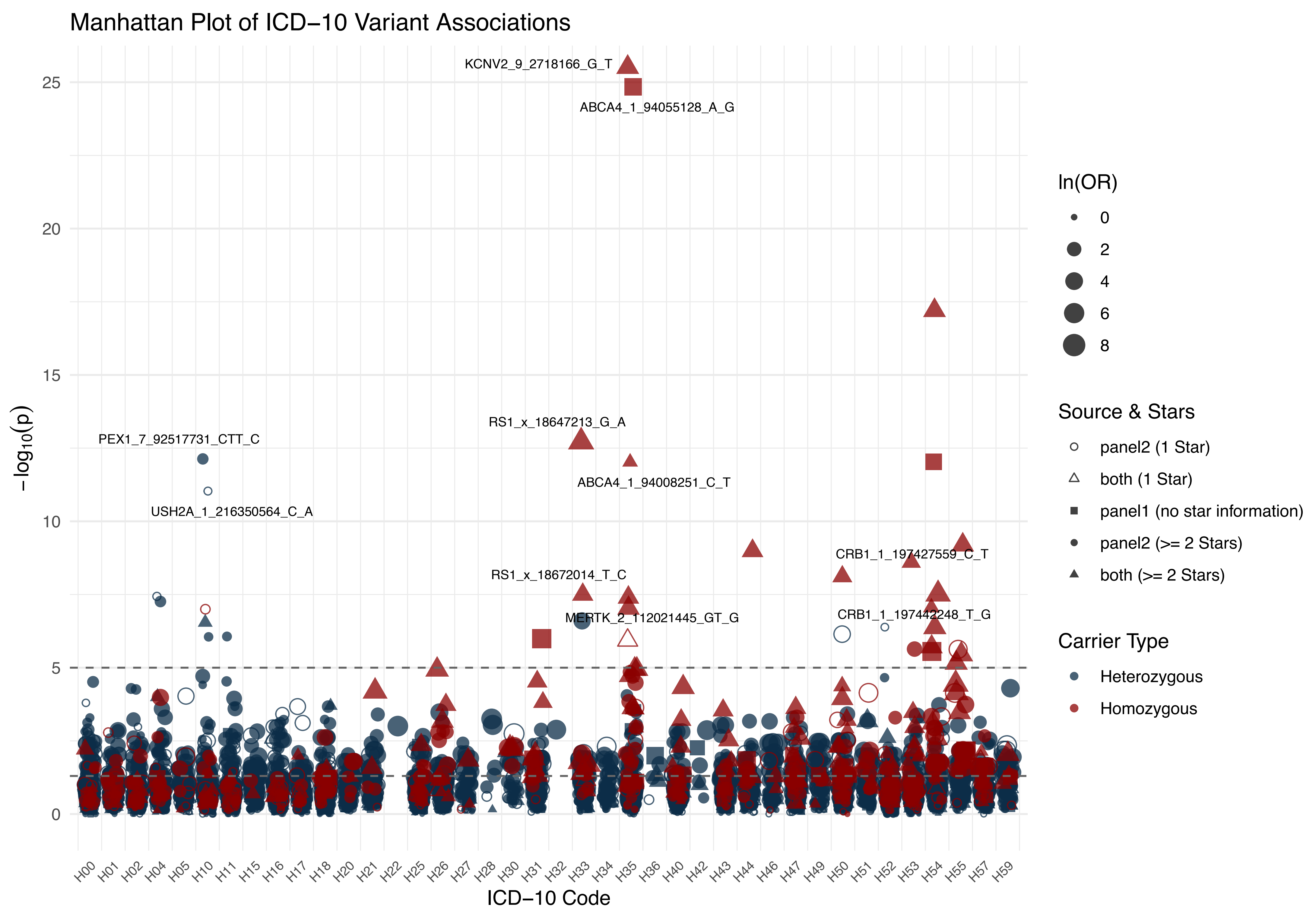

Manhattan Plot of ICD−10 Variant Associations
$-\log_{10}(p)$
0
5
10
15
20
25
KCNV2_9_2718166_G_T
ABCA4_1_94055128_A_G
PEX1_7_92517731_CTT_C
USH2A_1_216350564_C_A
RS1_x_18647213_G_A
ABCA4_1_94008251_C_T
RS1_x_18672014_T_C
MERTK_2_112021445_GT_G
CRB1_1_197427559_C_T
CRB1_1_197442248_T_G
H00 H01 H02 H04 H05 H10 H11 H15 H16 H17 H18 H20 H21 H22 H25 H26 H27 H28 H30 H31 H32 H33 H34 H35 H36 H40 H42 H43 H44 H46 H47 H49 H50 H51 H52 H53 H54 H55 H57 H59
ICD−10 Code
ln(OR)
0
2
4
6
8
Source & Stars
panel2 (1 Star)
both (1 Star)
panel1 (no star information)
panel2 (>= 2 Stars)
both (>= 2 Stars)
Carrier Type
Heterozygous
Homozygous

**Supplementary Figure 3. Manhattan plot of ICD-10-wide variant associations for inherited retinal disease (IRD) panel variants.**

Analyses were performed in the subset of 426,382 individuals with complete genomic and EHR data. After filtering to retain individual ICD-10 codes with a prevalence >1% in the EGP cohort, a total of 2,323 ICD-10 codes were analyzed across all subchapters. Each point represents a variant–ICD-10 subchapter association, plotted by the ICD-10 code (x-axis) and $-\log_{10}$(p-value) (y-axis). Heterozygous associations are shown in blue and homozygous in dark red, with point sizes reflecting the natural logarithm of the odds ratio (ln(OR)). Shapes denote variant source and ClinVar star status: filled shapes indicate ≥2-star submissions; unfilled shapes indicate 1-star entries. Variants with strong associations ($-\log_{10}p > 5$) are labeled with gene and variant identifiers. The strongest and biologically meaningful signals were observed for subchapters H35 (retinal disorders), H54 (visual impairment), H55 (nystagmus), H50 (strabismus), and H53 (visual disturbances), which were selected for downstream penetrance and burden estimation.

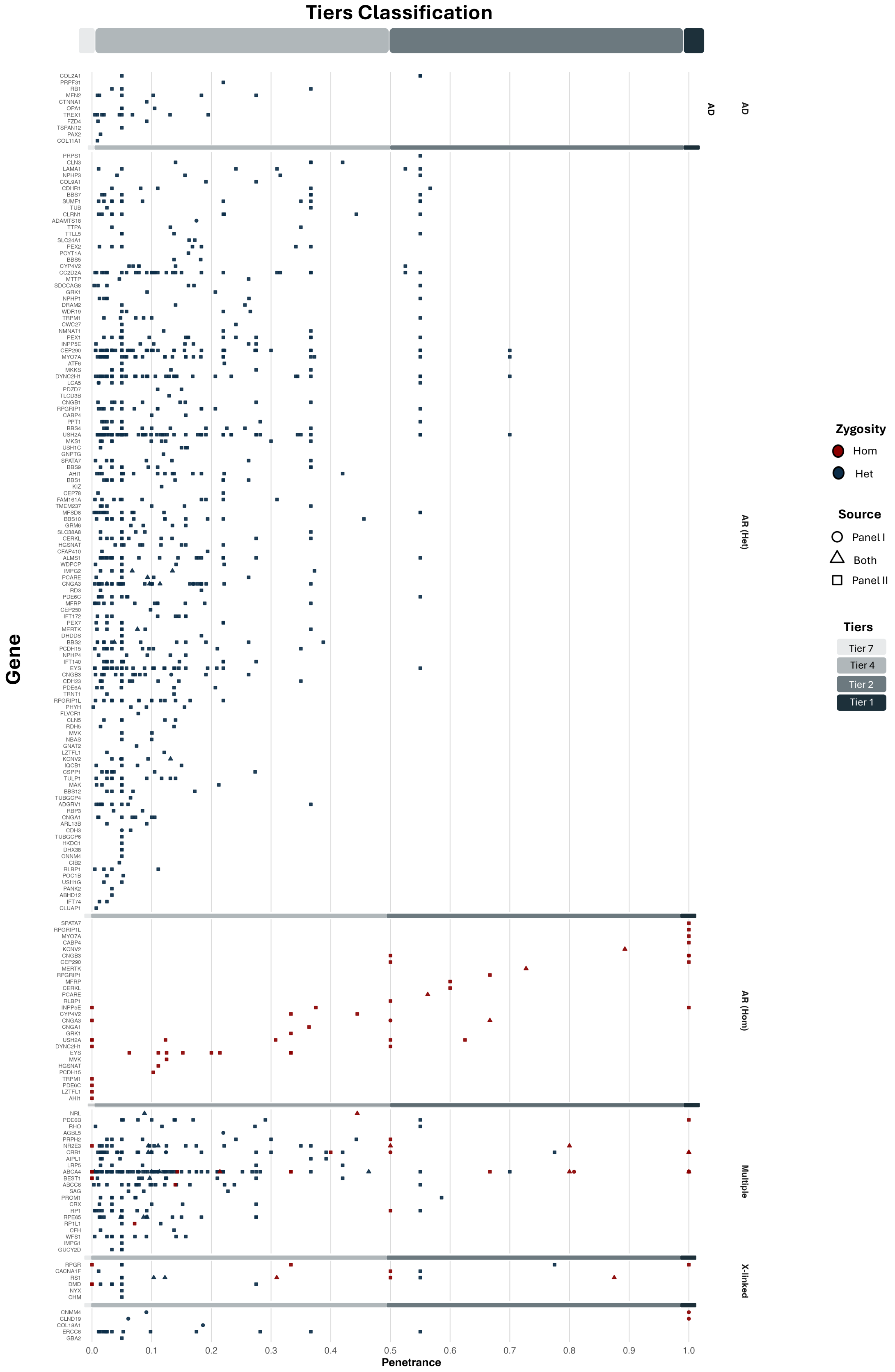
Tiers Classification
Gene
Penetrance
0.0
0.1
0.2
0.3
0.4
0.5
0.6
0.7
0.8
0.9
1.0
AD
AR (Het)
AR (Hom)
Multiple
X-linked
Zygosity
Hom
Het
Source
Panel I
Both
Panel II
Tiers
Tier 7
Tier 4
Tier 2
Tier 1

**Supplementary Figure 4. Penetrance of Pathogenic IRD Variants by Gene, Zygosity, and Mode of Inheritance (Including AR Heterozygotes).**

This plot summarizes variant-level penetrance estimates for 372 pathogenic and likely pathogenic variants in RetNet genes, stratified by mode of inheritance. Each point represents a variant, with penetrance calculated as the proportion of carriers presenting with any of the top five IRD-related ICD-10 phenotypes (Top5 Phenotypes: H35, H54, H55, H50, H53). Red symbols denote homozygous penetrance, and blue symbols denote heterozygous penetrance. Variant origin is indicated by shape (circle: Panel I, square: Panel II, triangle: both). Variants are annotated by penetrance tier: Tier 1 (fully penetrant, ≥2 carriers), Tier 2 (high penetrance ≥0.5, ≥2 carriers), Tier 4 (low penetrance <0.5, ≥2 carriers), and Tier 7 (significantly null, penetrance = 0, ≥2 carriers). Tiers requiring singleton carriers (Tiers 3, 5, 6) are excluded, as penetrance estimates based on single carriers were removed. For clarity, penetrance of autosomal recessive (AR) variants is shown separately for homozygous and heterozygous carriers. Within each inheritance mode, genes are ordered from highest to lowest mean penetrance.

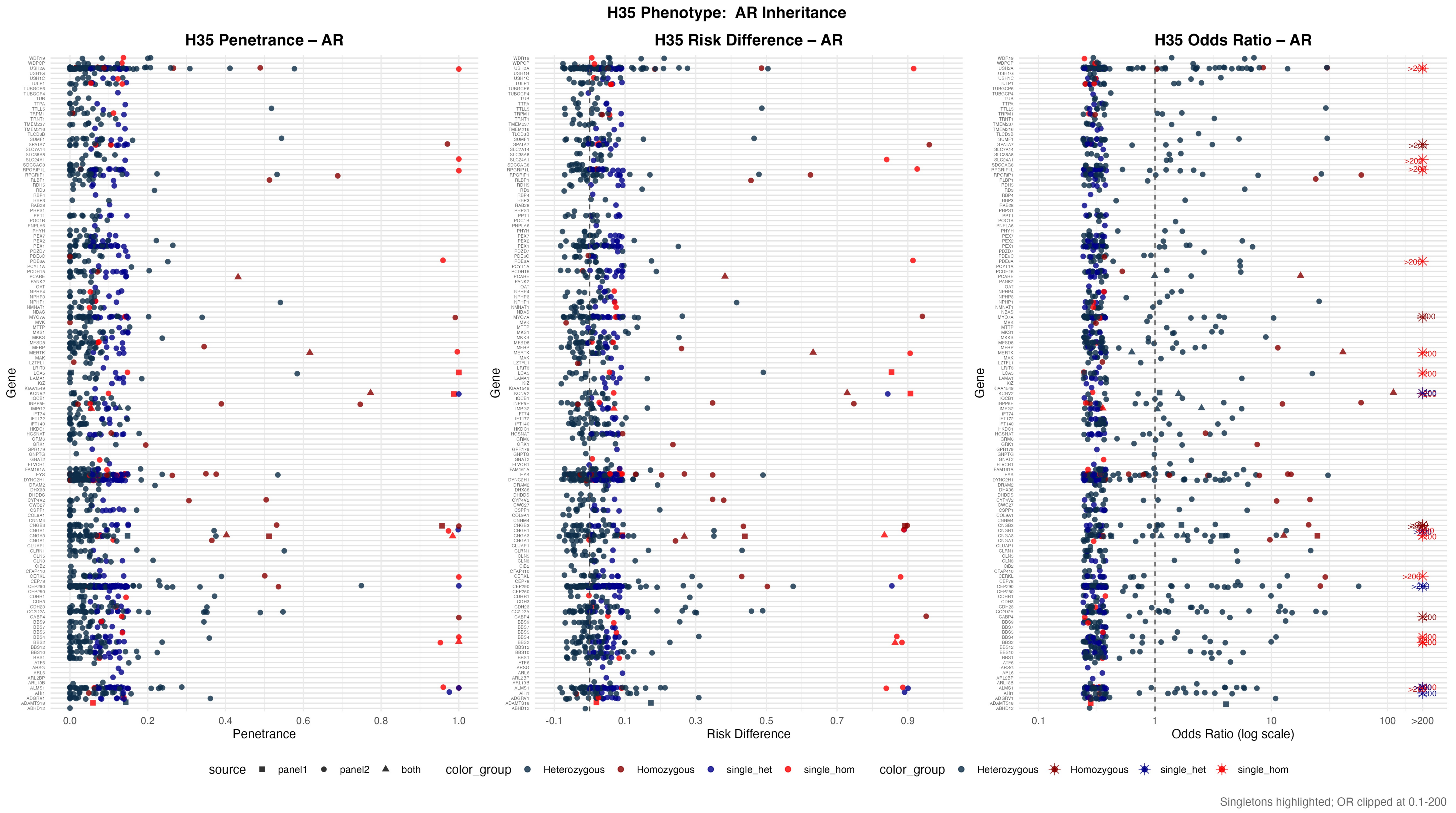

H35 Phenotype: AR Inheritance
H35 Penetrance – AR
H35 Risk Difference – AR
H35 Odds Ratio – AR
Gene
Penetrance
0.0
0.2
0.4
0.6
0.8
1.0
Risk Difference
-0.1
0.1
0.3
0.5
0.7
0.9
Odds Ratio (log scale)
0.1
1
10
100
>200
source
panel1
panel2
both
color_group
Heterozygous
Homozygous
single_het
single_hom
Singletons highlighted; OR clipped at 0.1-200

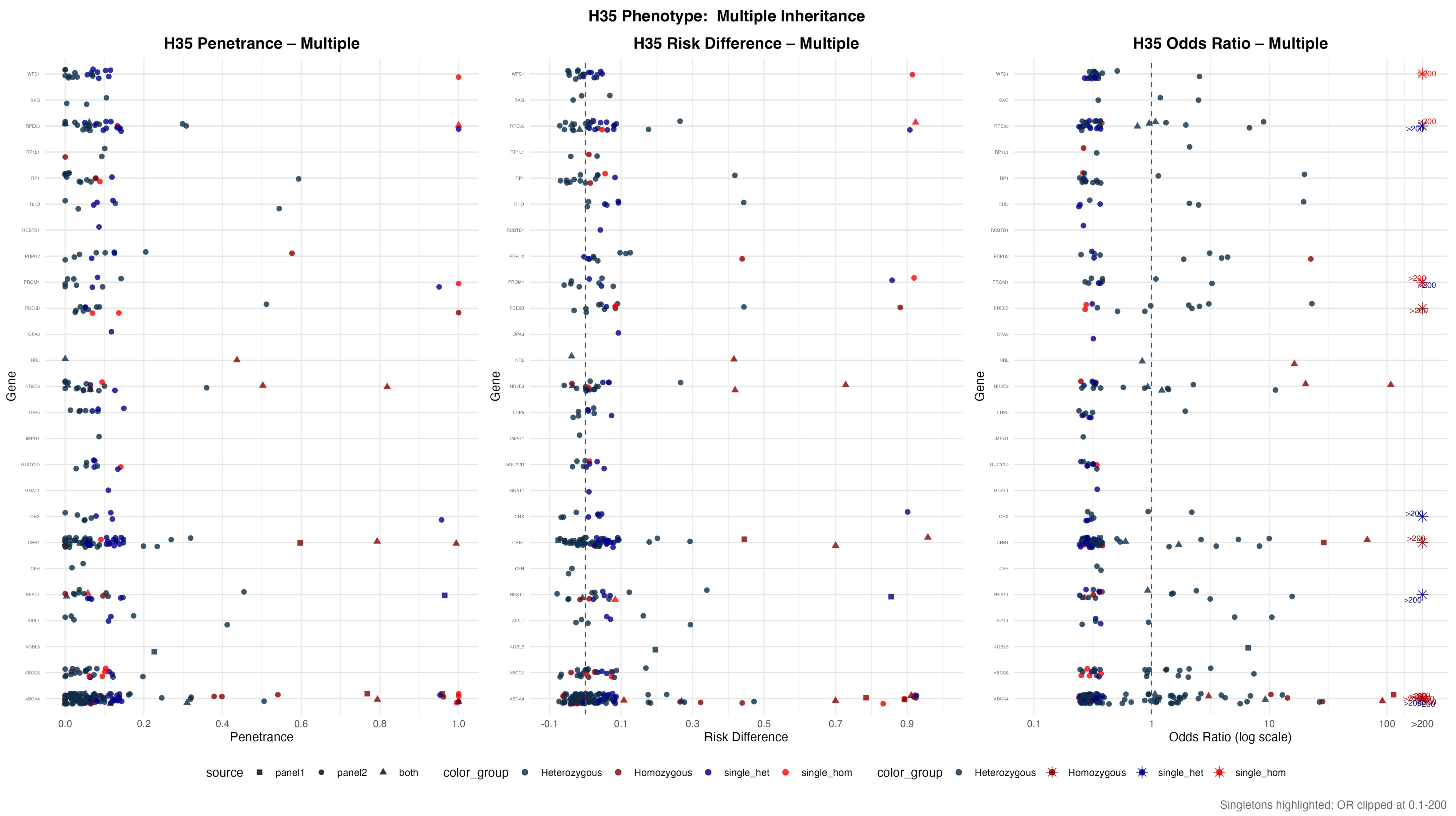
H35 Phenotype: Multiple Inheritance
H35 Penetrance – Multiple
H35 Risk Difference – Multiple
H35 Odds Ratio – Multiple
Gene
WFS1
SAG
RPE65
RP1L1
RP1
RHO
RCBTB1
PRPH2
PROM1
PDE6B
OPA3
NRL
NR2E3
LRP5
IMPG1
GUCY2D
GNAT1
CRX
CRB1
CFH
BEST1
AIPL1
AGBL5
ABCC6
ABCA4
Penetrance
Risk Difference
Odds Ratio (log scale)
source
panel1
panel2
both
color_group
Heterozygous
Homozygous
single_het
single_hom
Singletons highlighted; OR clipped at 0.1-200

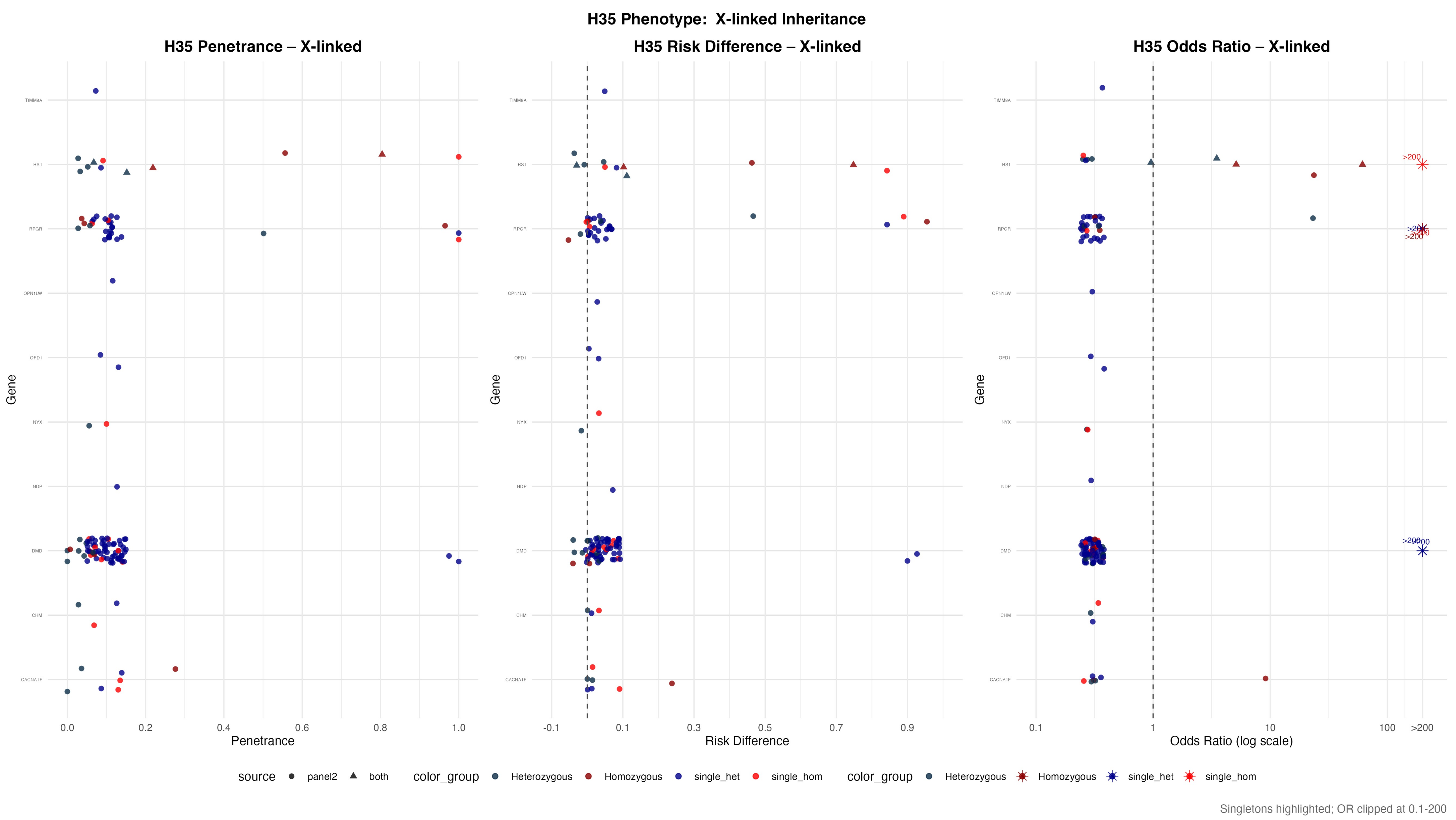
H35 Phenotype: X-linked Inheritance
H35 Penetrance – X-linked
H35 Risk Difference – X-linked
H35 Odds Ratio – X-linked
Gene
TIMM8A
RS1
RPGR
OPN1LW
OFD1
NYX
NDP
DMD
CHM
CACNA1F
0.0
0.2
0.4
0.6
0.8
1.0
Penetrance
-0.1
0.1
0.3
0.5
0.7
0.9
Risk Difference
0.1
1
10
100
>200
Odds Ratio (log scale)
source
panel2
both
color_group
Heterozygous
Homozygous
single_het
single_hom
Singletons highlighted; OR clipped at 0.1-200

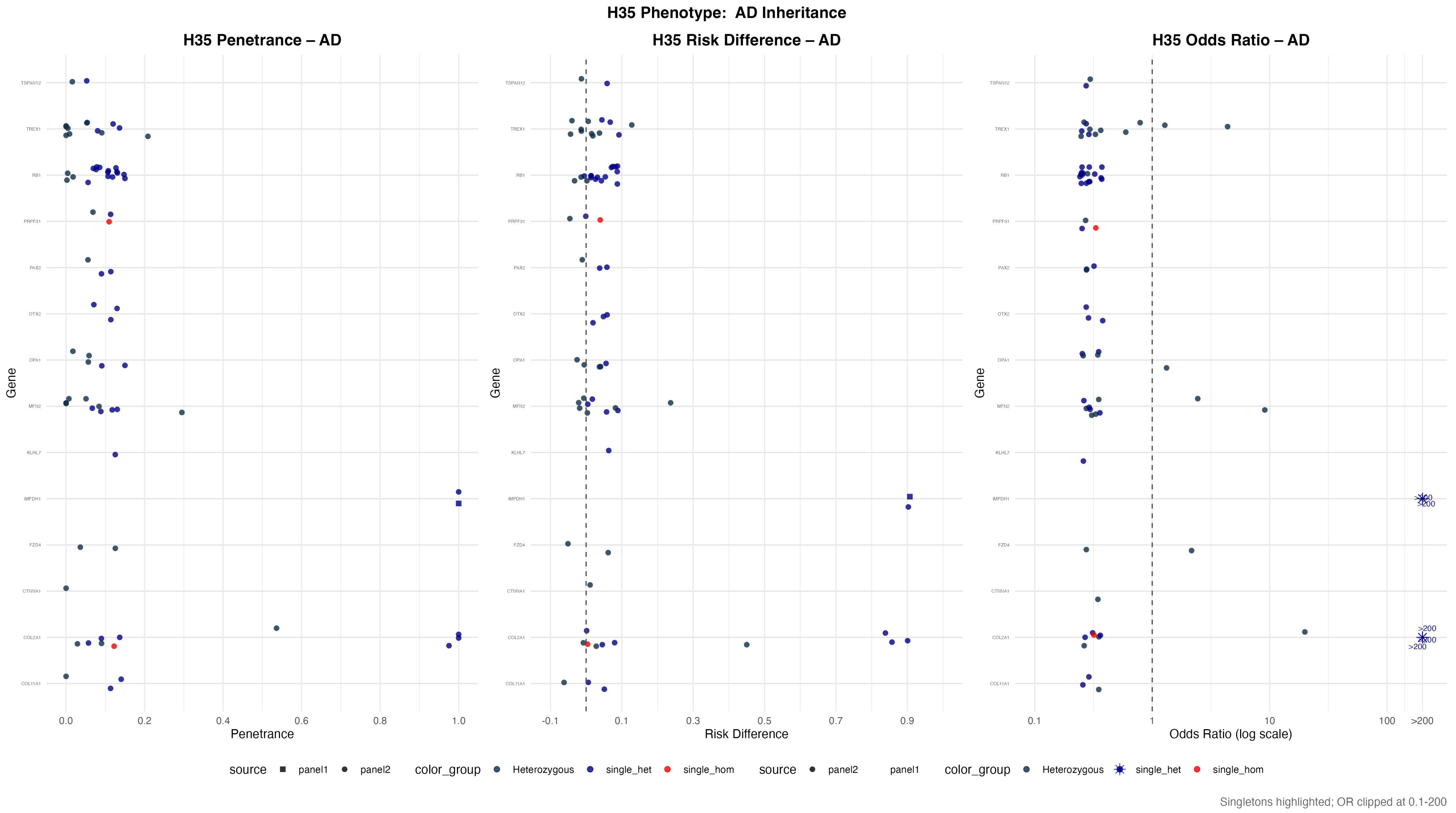

H35 Phenotype: AD Inheritance
H35 Penetrance – AD
H35 Risk Difference – AD
H35 Odds Ratio – AD
Gene
TSPAN12
TREX1
RB1
PRPF31
PAX2
OTX2
OPA1
MFN2
KLHL7
IMPDH1
FZD4
CTNNA1
COL2A1
COL11A1
Penetrance
Risk Difference
Odds Ratio (log scale)
>200
source
panel1
panel2
color_group
Heterozygous
single_het
single_hom
Singletons highlighted; OR clipped at 0.1-200

**Supplementary Figure 6. Variant-Level Penetrance and Association Metrics for Retinal Disorders (H35 Subchapter).**

Estimated clinical penetrance (left), risk difference (center), and odds ratio (right) are shown for heterozygous and homozygous carriers of known IRD-associated variants, aggregated by gene and stratified by inheritance mode: (A) autosomal recessive (AR), (B) autosomal dominant (AD), (C) multiple inheritance patterns, and (D) X-linked genes. Each point represents a variant with linked genotype and phenotype data from 81,080 carriers. Outcomes are defined as presence of at least one ICD-10 code within subchapter H35 (retinal disorders). Colors and shapes denote zygosity and variant source (Panel I vs. Panel II). Vertical dashed lines indicate the null values for risk difference (0) and odds ratio (1). Variants with extreme OR values are plotted at axis limits for clarity. This subchapter-specific analysis refines our understanding of variant expressivity by focusing on the most directly relevant diagnostic category for inherited retinal disorders.

### H50 Phenotype: X-linked Inheritance

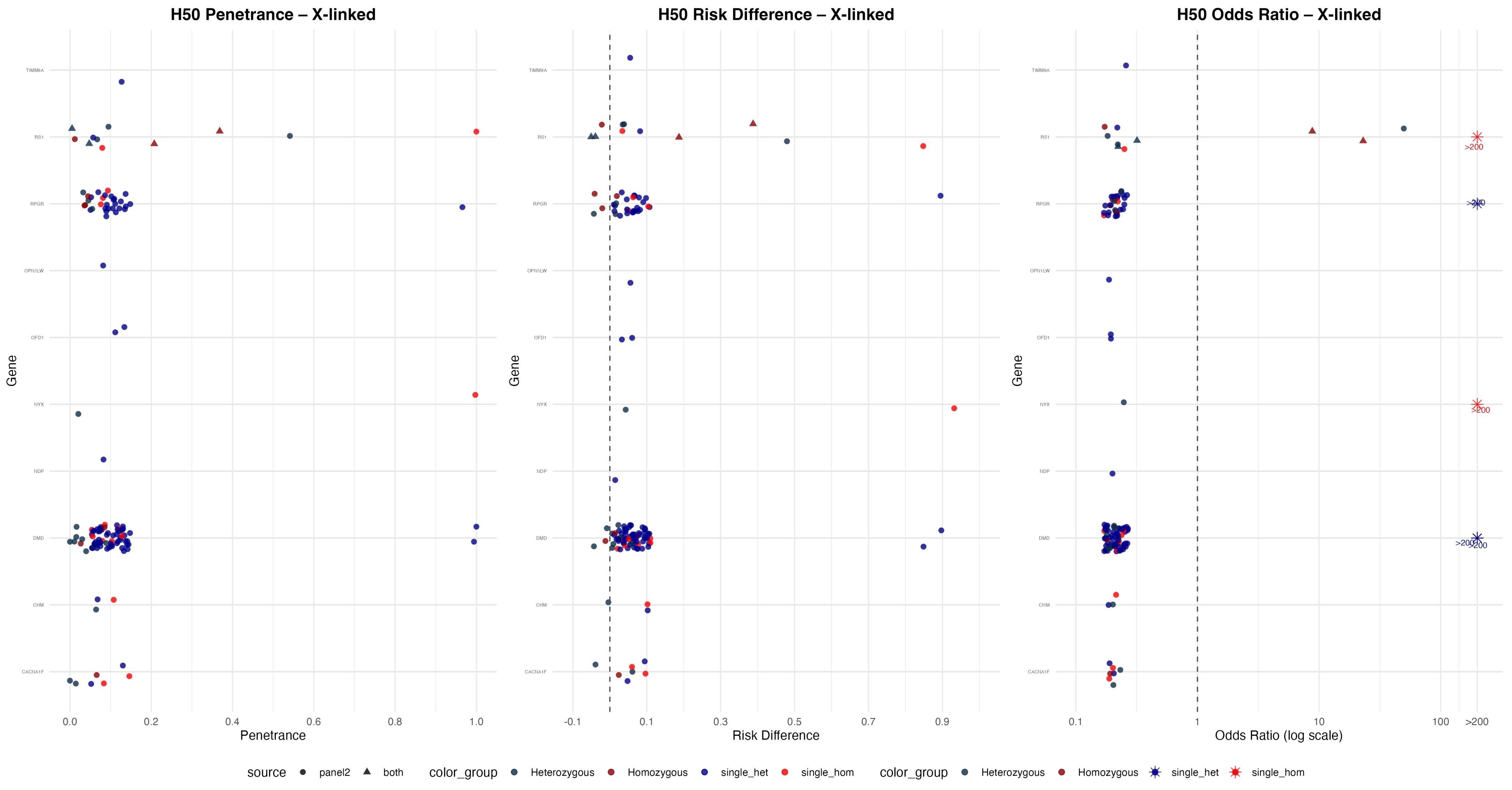


Singletons highlighted; OR clipped at 0.1-200

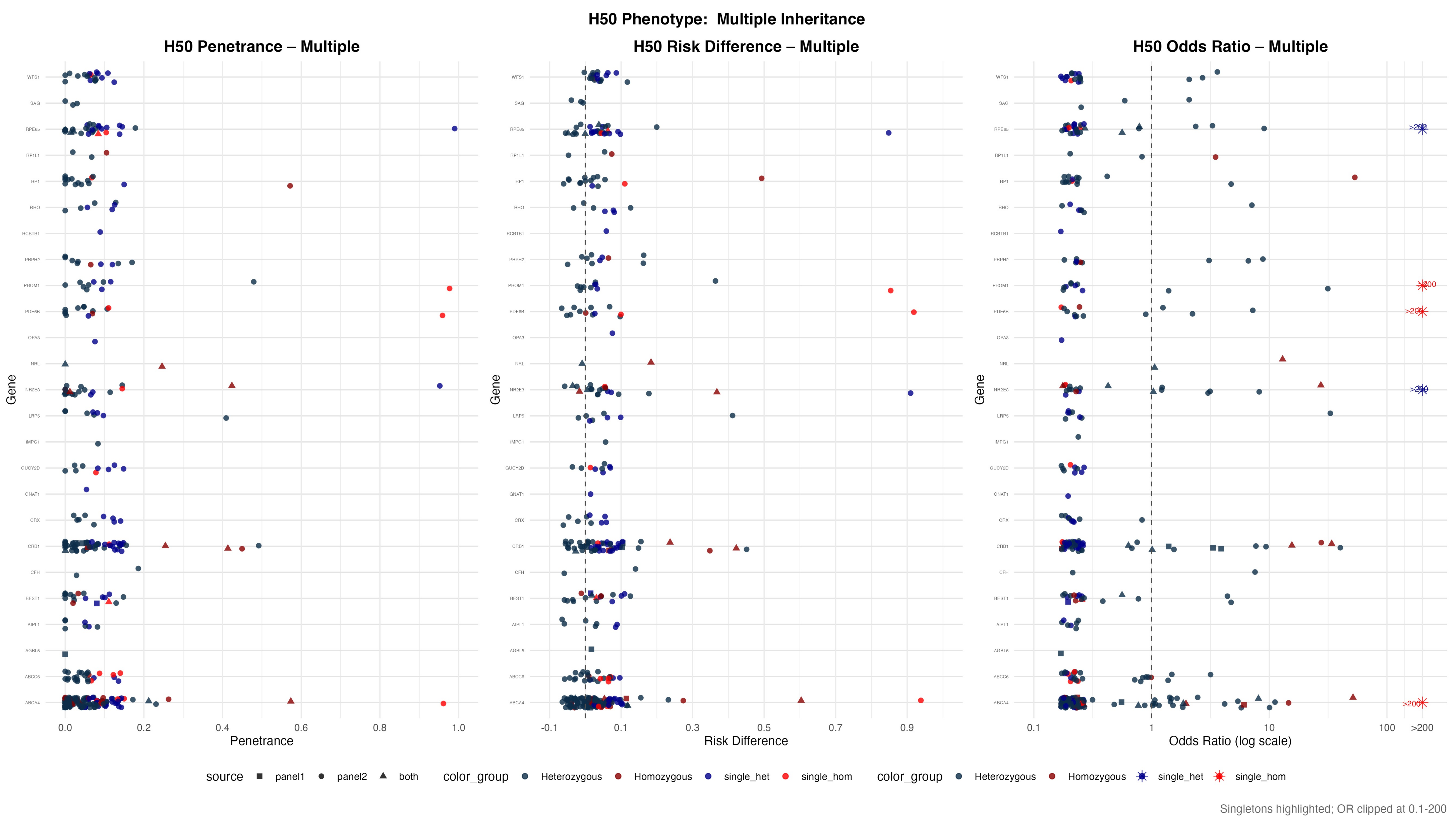
H50 Phenotype: Multiple Inheritance
H50 Penetrance – Multiple
H50 Risk Difference – Multiple
H50 Odds Ratio – Multiple
Gene
WFS1
SAG
RPE65
RP1L1
RP1
RHO
RCBTB1
PRPH2
PROM1
PDE6B
OPA3
NRL
NR2E3
LRP5
IMPG1
GUCY2D
GNAT1
CRX
CRB1
CFH
BEST1
AIPL1
AGBL5
ABCC6
ABCA4
Penetrance
Risk Difference
Odds Ratio (log scale)
>200
source
panel1
panel2
both
color_group
Heterozygous
Homozygous
single_het
single_hom
Singletons highlighted; OR clipped at 0.1-200

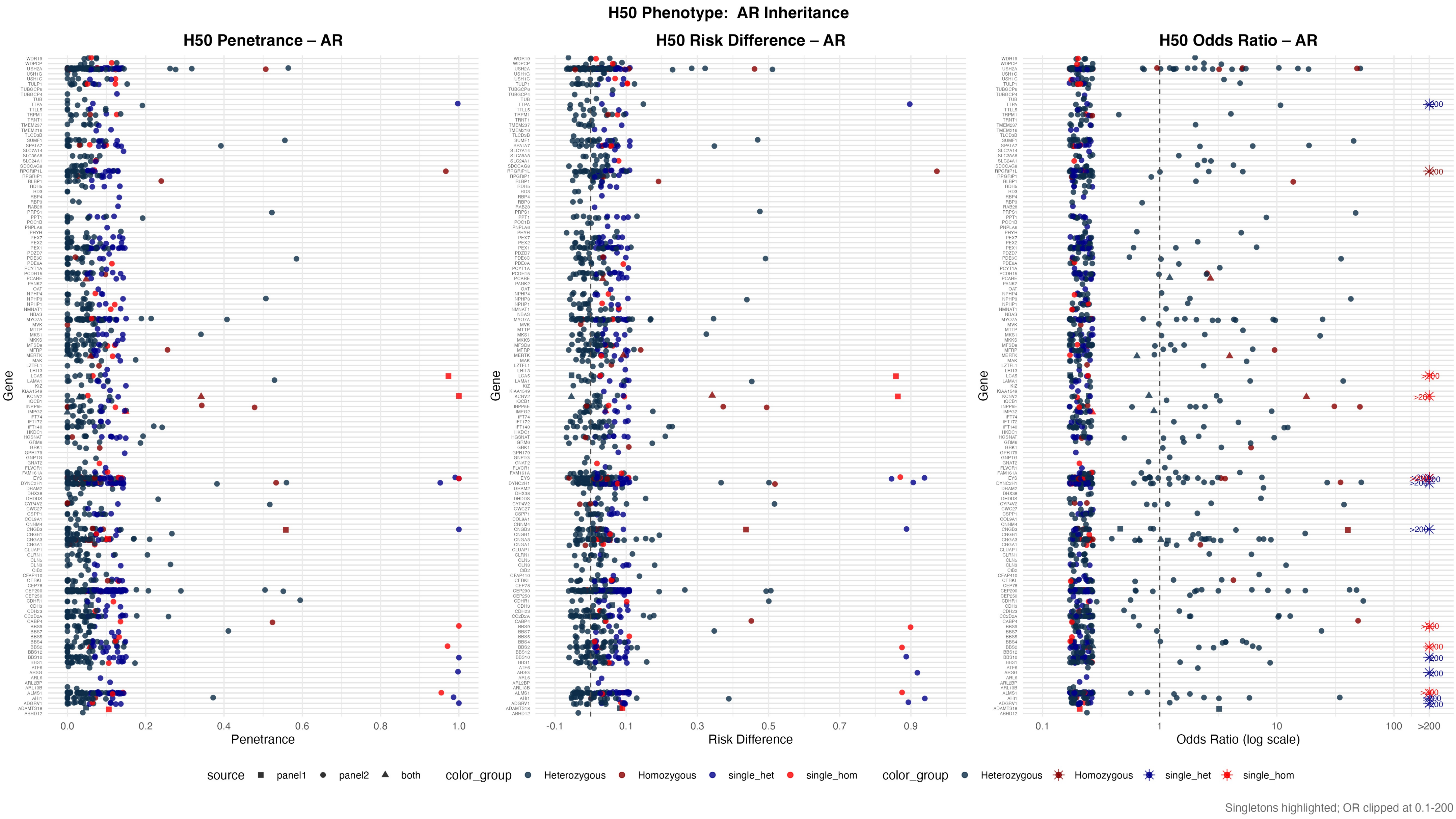
H50 Phenotype: AR Inheritance
H50 Penetrance – AR
H50 Risk Difference – AR
H50 Odds Ratio – AR
Gene
Penetrance
Risk Difference
Odds Ratio (log scale)
source panel1 panel2 both
color_group Heterozygous Homozygous single_het single_hom
Singletons highlighted; OR clipped at 0.1-200

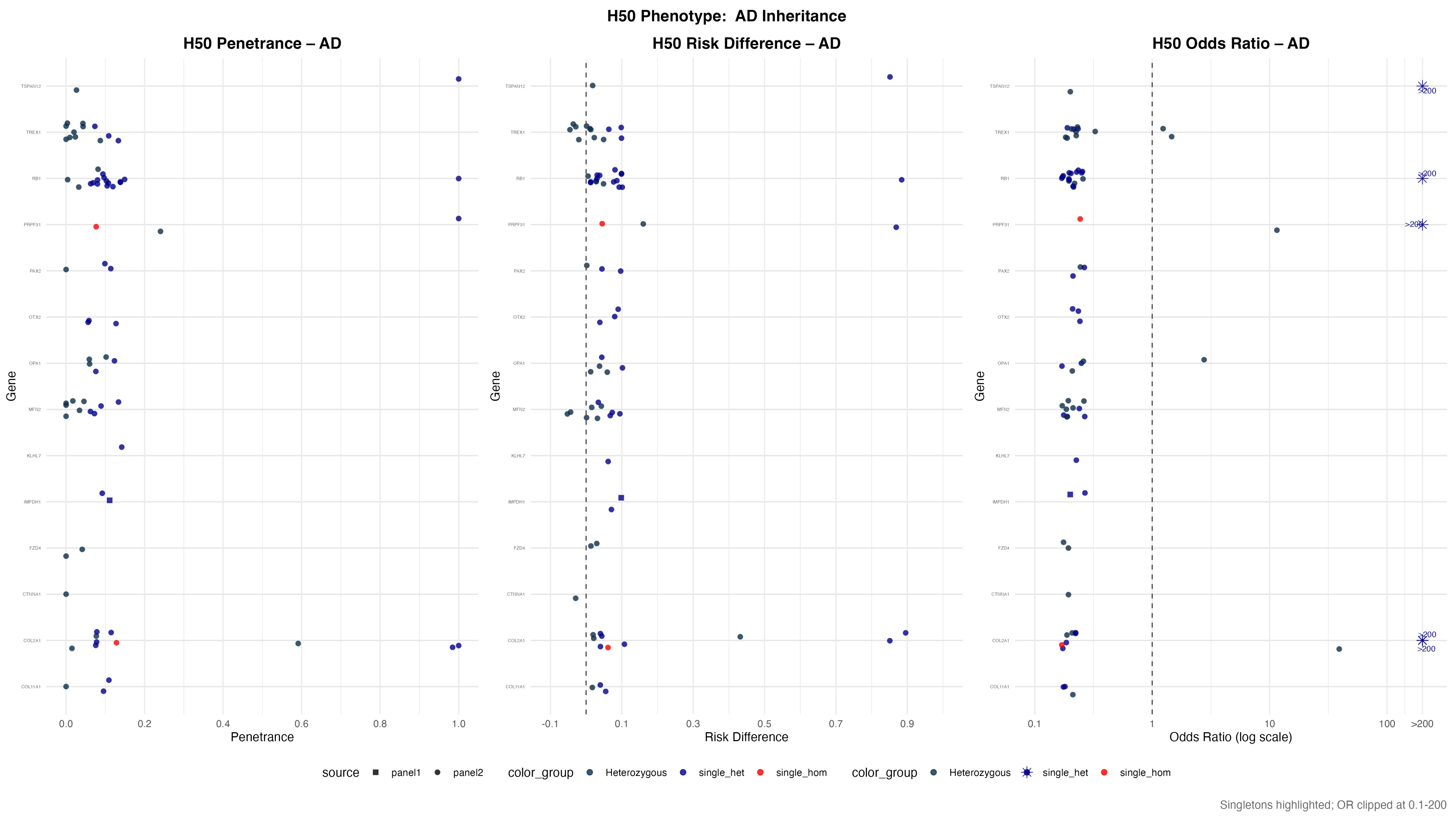

H50 Phenotype: AD Inheritance
H50 Penetrance – AD
H50 Risk Difference – AD
H50 Odds Ratio – AD
Gene
TSPAN12
TREX1
RB1
PRPF31
PAX2
OTX2
OPA1
MFN2
KLHL7
IMPDH1
FZD4
CTNNA1
COL2A1
COL11A1
Penetrance
Risk Difference
Odds Ratio (log scale)
>200
source
panel1
panel2
color_group
Heterozygous
single_het
single_hom
Singletons highlighted; OR clipped at 0.1-200

**Supplementary Figure 7. Variant-Level Penetrance and Association Metrics for Retinal Disorders (H50 Subchapter).**

Estimated clinical penetrance (left), risk difference (center), and odds ratio (right) are shown for heterozygous and homozygous carriers of known IRD-associated variants, aggregated by gene and stratified by inheritance mode: (A) autosomal recessive (AR), (B) autosomal dominant (AD), (C) multiple inheritance patterns, and (D) X-linked genes. Each point represents a variant with linked genotype and phenotype data from 81,080 carriers. Outcomes are defined as presence of at least one ICD-10 code within subchapter H50 (strabismus). Colors and shapes denote zygosity and variant source (Panel I vs. Panel II). Vertical dashed lines indicate the null values for risk difference (0) and odds ratio (1). Variants with extreme OR values are plotted at axis limits for clarity. This subchapter-specific analysis helps clarify which IRD variants contribute to the clinical manifestation of ocular misalignment and motility disorders.

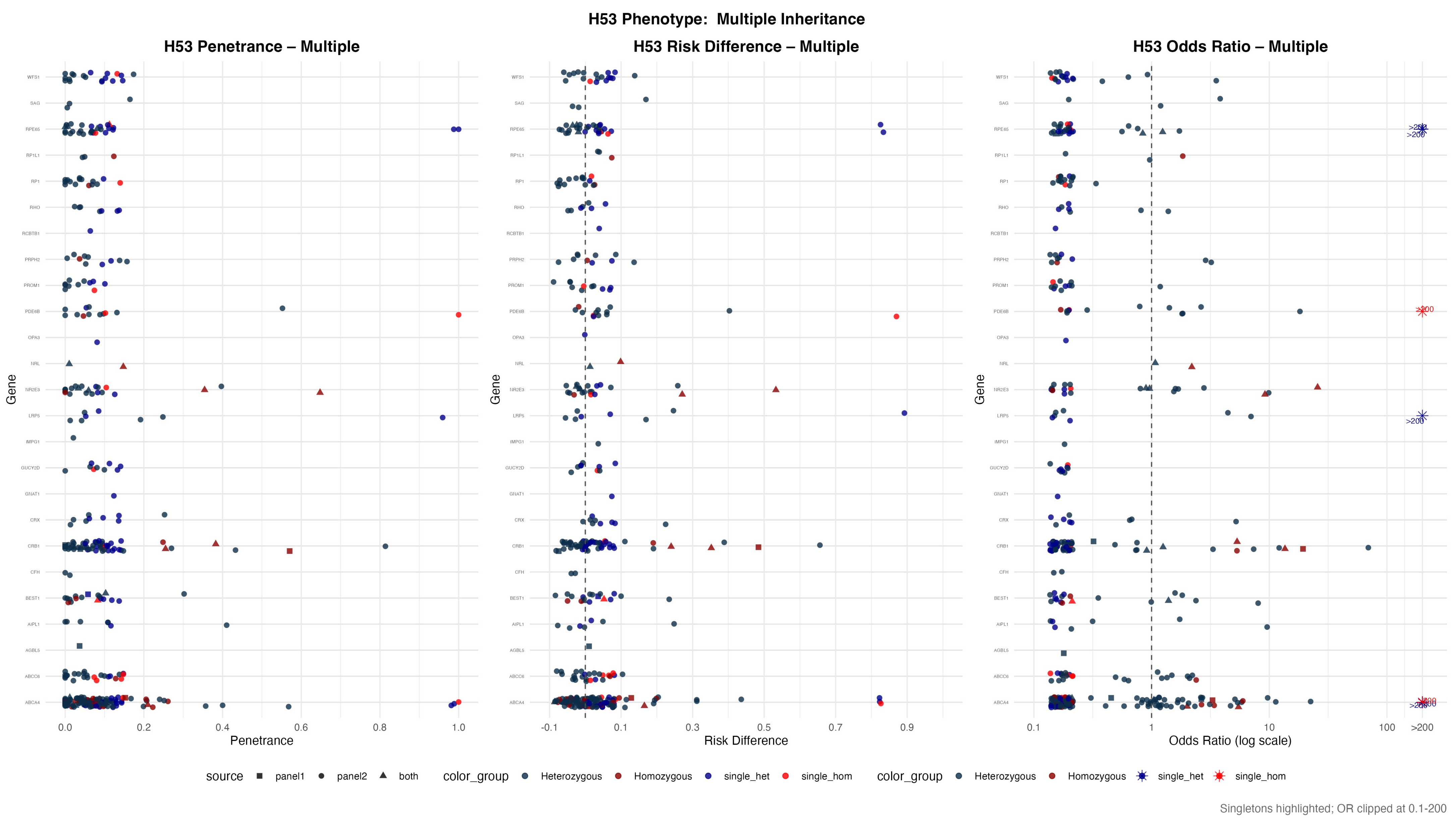

H53 Phenotype: Multiple Inheritance
H53 Penetrance – Multiple
H53 Risk Difference – Multiple
H53 Odds Ratio – Multiple
Gene
WFS1
SAG
RPE65
RP1L1
RP1
RHO
RCBTB1
PRPH2
PROM1
PDE6B
OPA3
NRL
NR2E3
LRP5
IMPG1
GUCY2D
GNAT1
CRX
CRB1
CFH
BEST1
AIPL1
AGBL5
ABCC6
ABCA4
Penetrance
Risk Difference
Odds Ratio (log scale)
>200
source
panel1
panel2
both
color_group
Heterozygous
Homozygous
single_het
single_hom
Singletons highlighted; OR clipped at 0.1-200

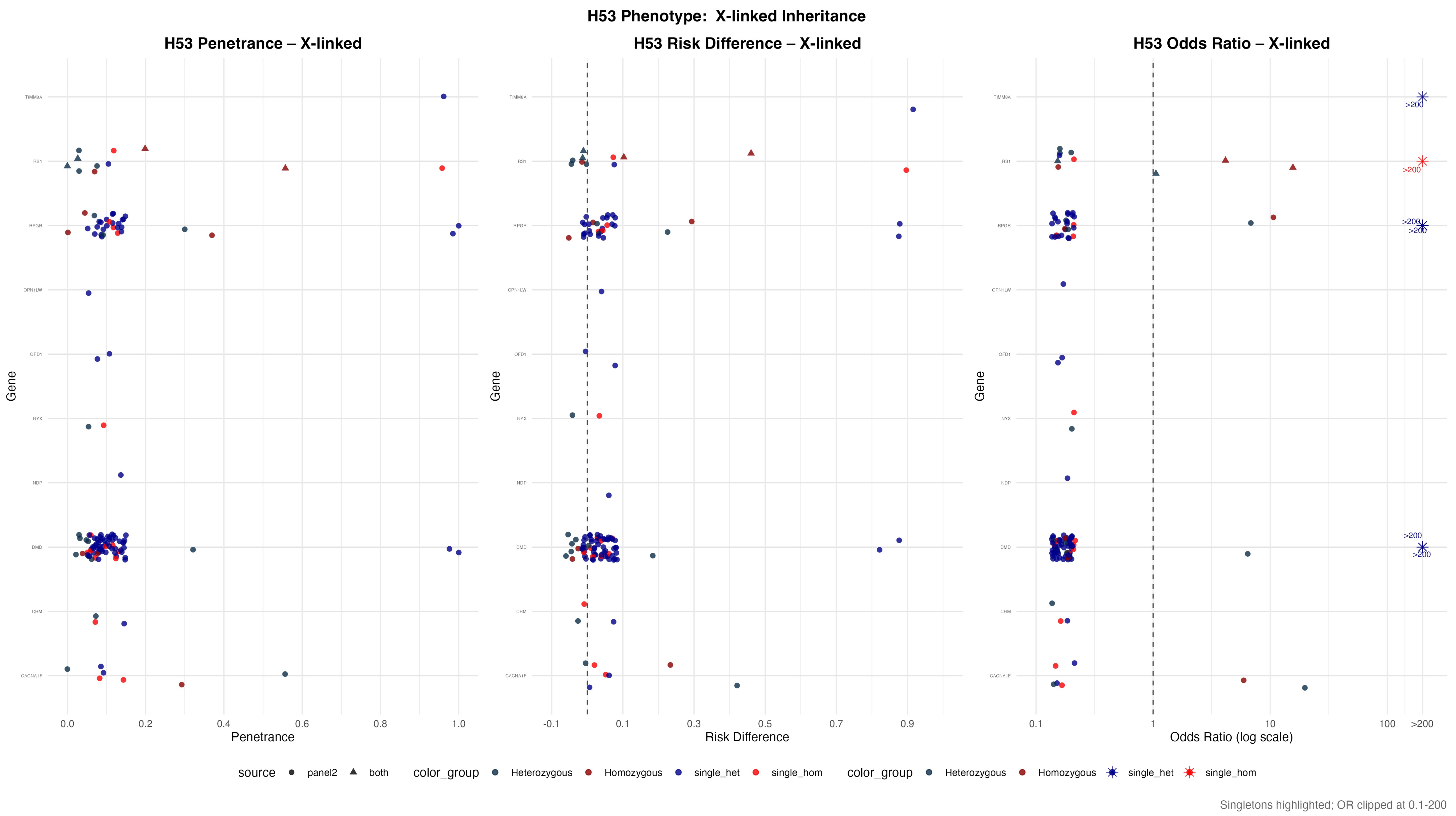
H53 Phenotype: X-linked Inheritance
H53 Penetrance – X-linked
H53 Risk Difference – X-linked
H53 Odds Ratio – X-linked
Gene
TIMM8A
RS1
RPGR
OPN1LW
OFD1
NYX
NDP
DMD
CHM
CACNA1F
0.0
0.2
0.4
0.6
0.8
1.0
Penetrance
-0.1
0.1
0.3
0.5
0.7
0.9
Risk Difference
0.1
1
10
100
>200
Odds Ratio (log scale)
source
panel2
both
color_group
Heterozygous
Homozygous
single_het
single_hom
Singletons highlighted; OR clipped at 0.1-200

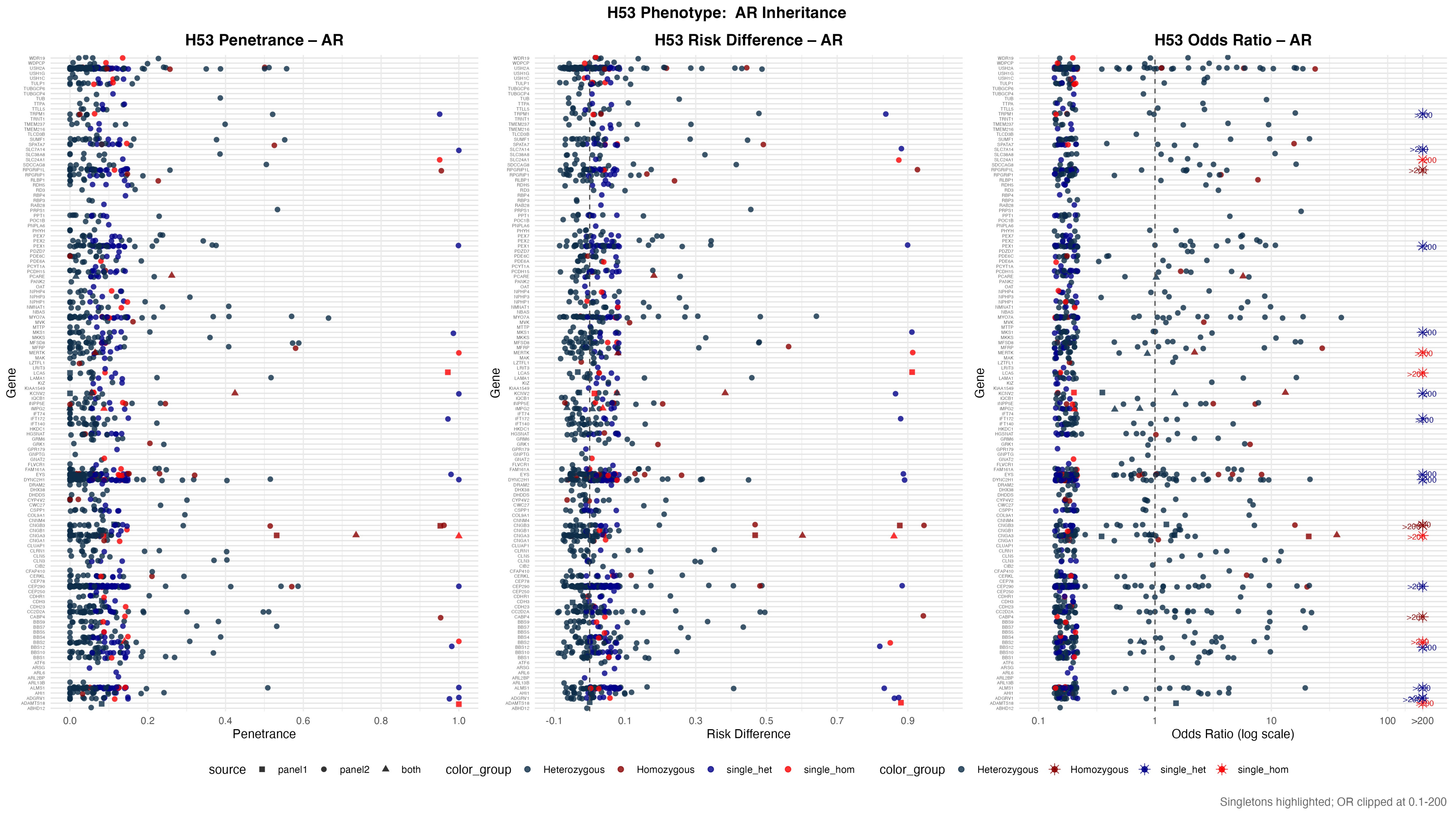

H53 Phenotype: AR Inheritance
H53 Penetrance – AR
H53 Risk Difference – AR
H53 Odds Ratio – AR
Gene
Penetrance
Risk Difference
Odds Ratio (log scale)
source
panel1
panel2
both
color_group
Heterozygous
Homozygous
single_het
single_hom
Singletons highlighted; OR clipped at 0.1-200

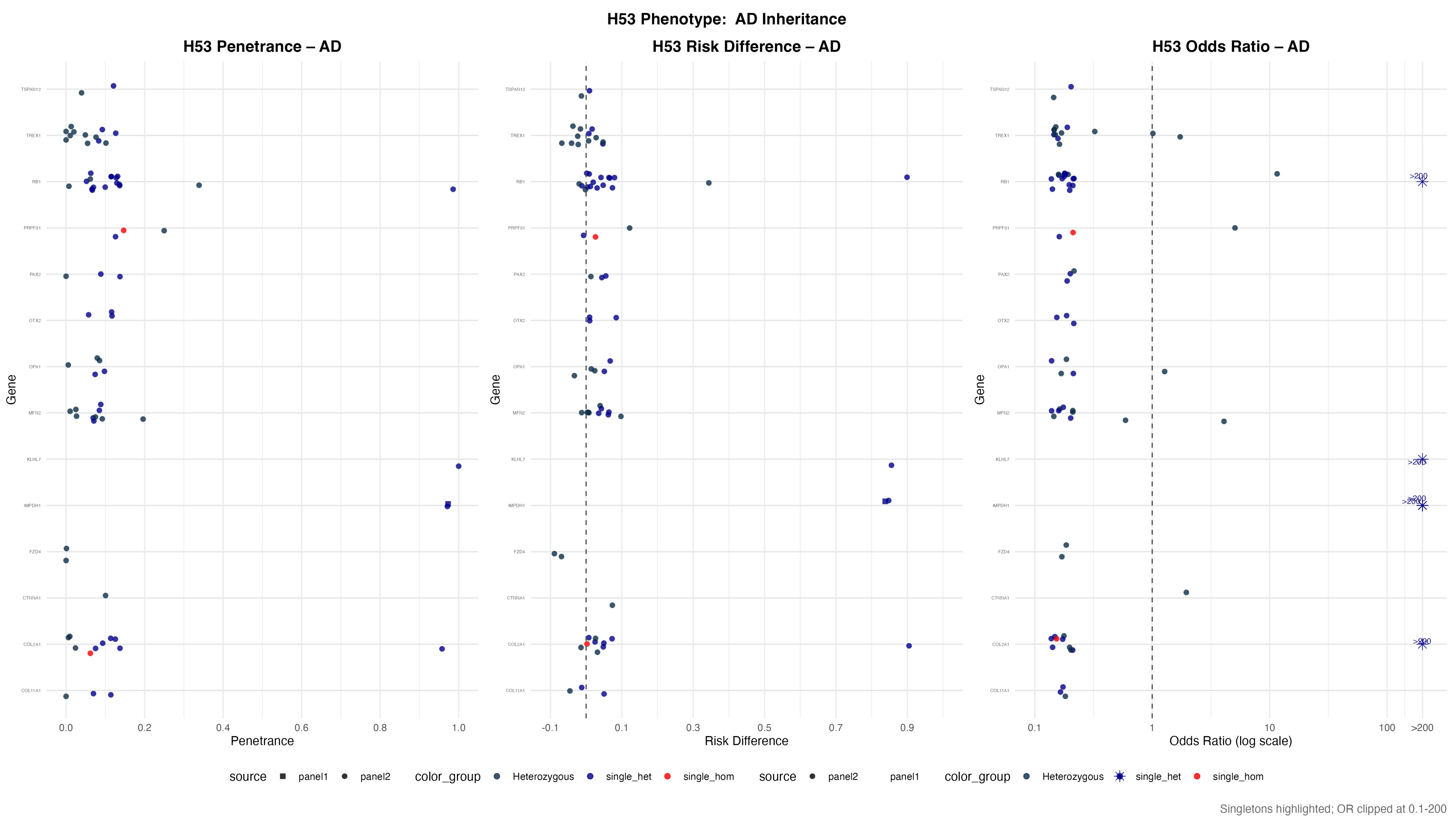
H53 Phenotype: AD Inheritance
H53 Penetrance – AD
H53 Risk Difference – AD
H53 Odds Ratio – AD
Gene
TSPAN12
TREX1
RB1
PRPF31
PAX2
OTX2
OPA1
MFN2
KLHL7
IMPDH1
FZD4
CTNNA1
COL2A1
COL11A1
0.0
0.2
0.4
0.6
0.8
1.0
Penetrance
-0.1
0.1
0.3
0.5
0.7
0.9
Risk Difference
0.1
1
10
100
>200
Odds Ratio (log scale)
source
panel1
panel2
color_group
Heterozygous
single_het
single_hom
Singletons highlighted; OR clipped at 0.1-200

**Supplementary Figure 8. Variant-Level Penetrance and Association Metrics for Retinal Disorders (H53 Subchapter).**

Estimated clinical penetrance (left), risk difference (center), and odds ratio (right) are shown for heterozygous and homozygous carriers of known IRD-associated variants, aggregated by gene and stratified by inheritance mode: (A) autosomal recessive (AR), (B) autosomal dominant (AD), (C) multiple inheritance patterns, and (D) X-linked genes. Each point represents a variant with linked genotype and phenotype data from 81,080 carriers. Outcomes are defined as presence of at least one ICD-10 code within subchapter H53 (rvisual disturbances). Colors and shapes denote zygosity and variant source (Panel I vs. Panel II). Vertical dashed lines indicate the null values for risk difference (0) and odds ratio (1). Variants with extreme OR values are plotted at axis limits for clarity. This subchapter-specific analysis highlights IRD variants contributing to early visual symptoms, such as photophobia, scotomas, and reduced visual acuity, supporting their clinical relevance even prior to end-stage vision loss.

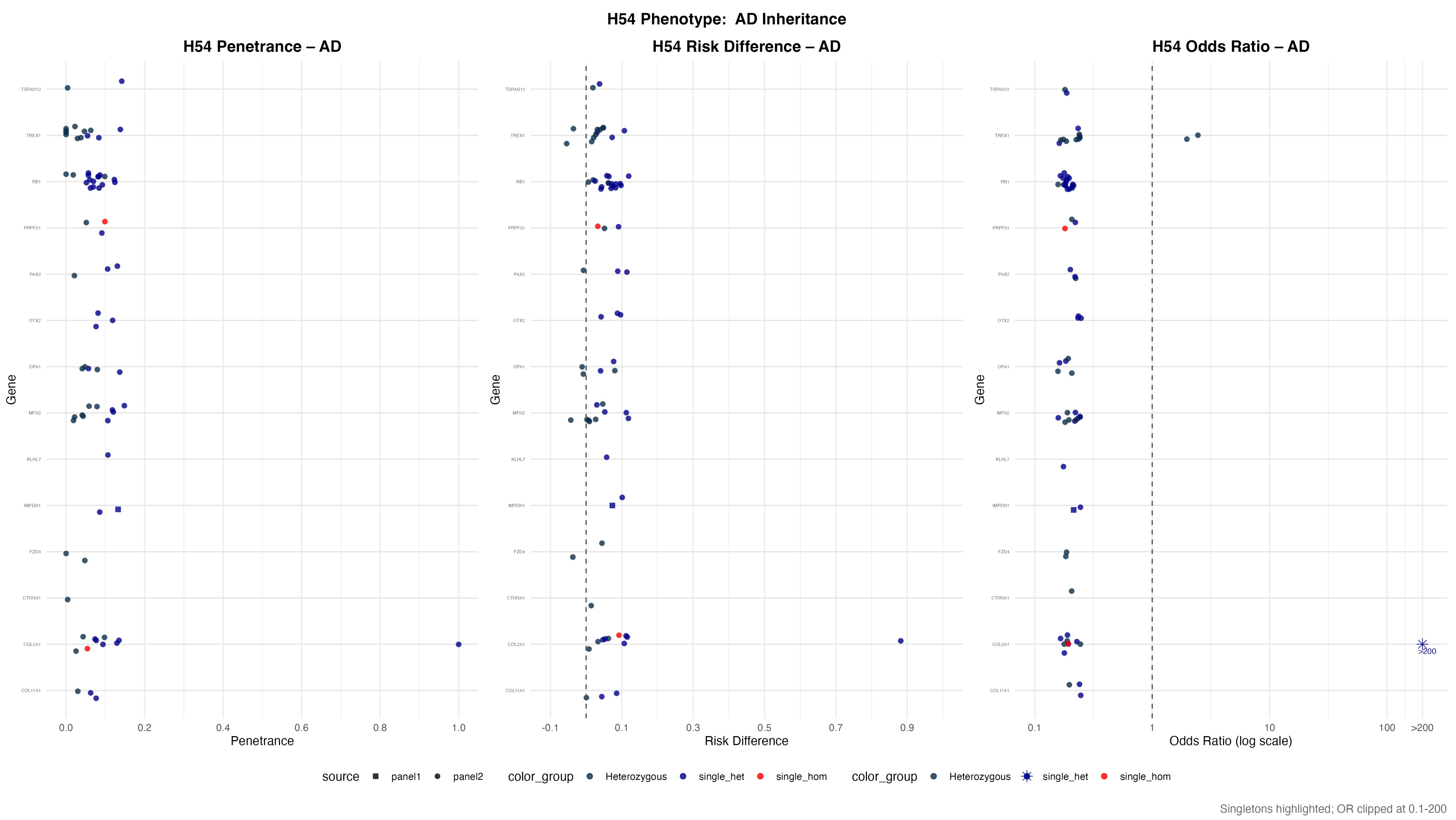
H54 Phenotype: AD Inheritance
H54 Penetrance – AD
H54 Risk Difference – AD
H54 Odds Ratio – AD
Gene
TSPAN12
TREX1
RB1
PRPF31
PAX2
OTX2
OPA1
MFN2
KLHL7
IMPDH1
FZD4
CTNNA1
COL2A1
COL11A1
0.0
0.2
0.4
0.6
0.8
1.0
Penetrance
-0.1
0.1
0.3
0.5
0.7
0.9
Risk Difference
0.1
1
10
100
>200
Odds Ratio (log scale)
source
panel1
panel2
color_group
Heterozygous
single_het
single_hom
Singletons highlighted; OR clipped at 0.1-200

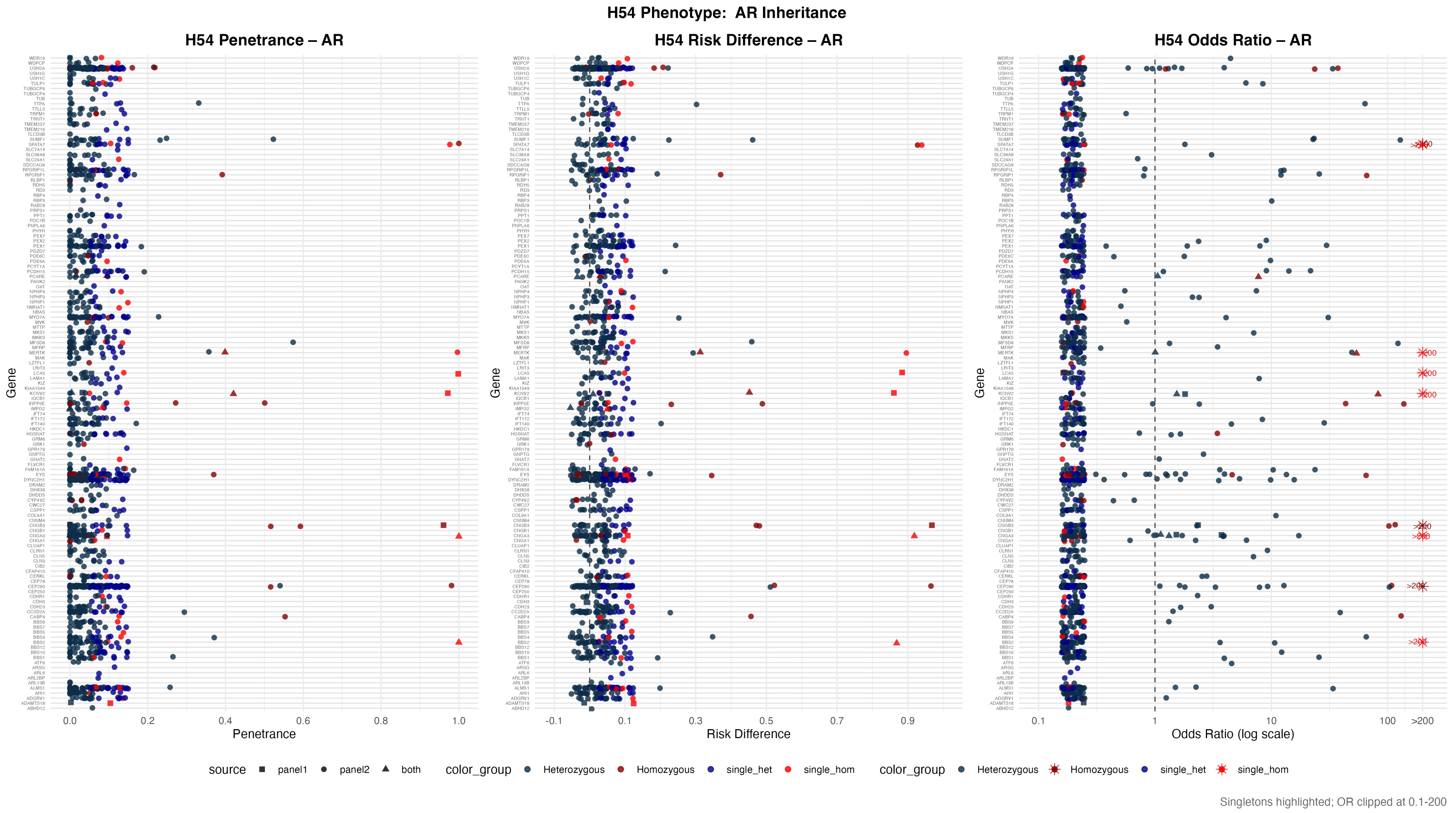
H54 Phenotype: AR Inheritance
H54 Penetrance – AR
H54 Risk Difference – AR
H54 Odds Ratio – AR
Gene
Penetrance
Risk Difference
Odds Ratio (log scale)
source
panel1
panel2
both
color_group
Heterozygous
Homozygous
single_het
single_hom
>200
Singletons highlighted; OR clipped at 0.1-200

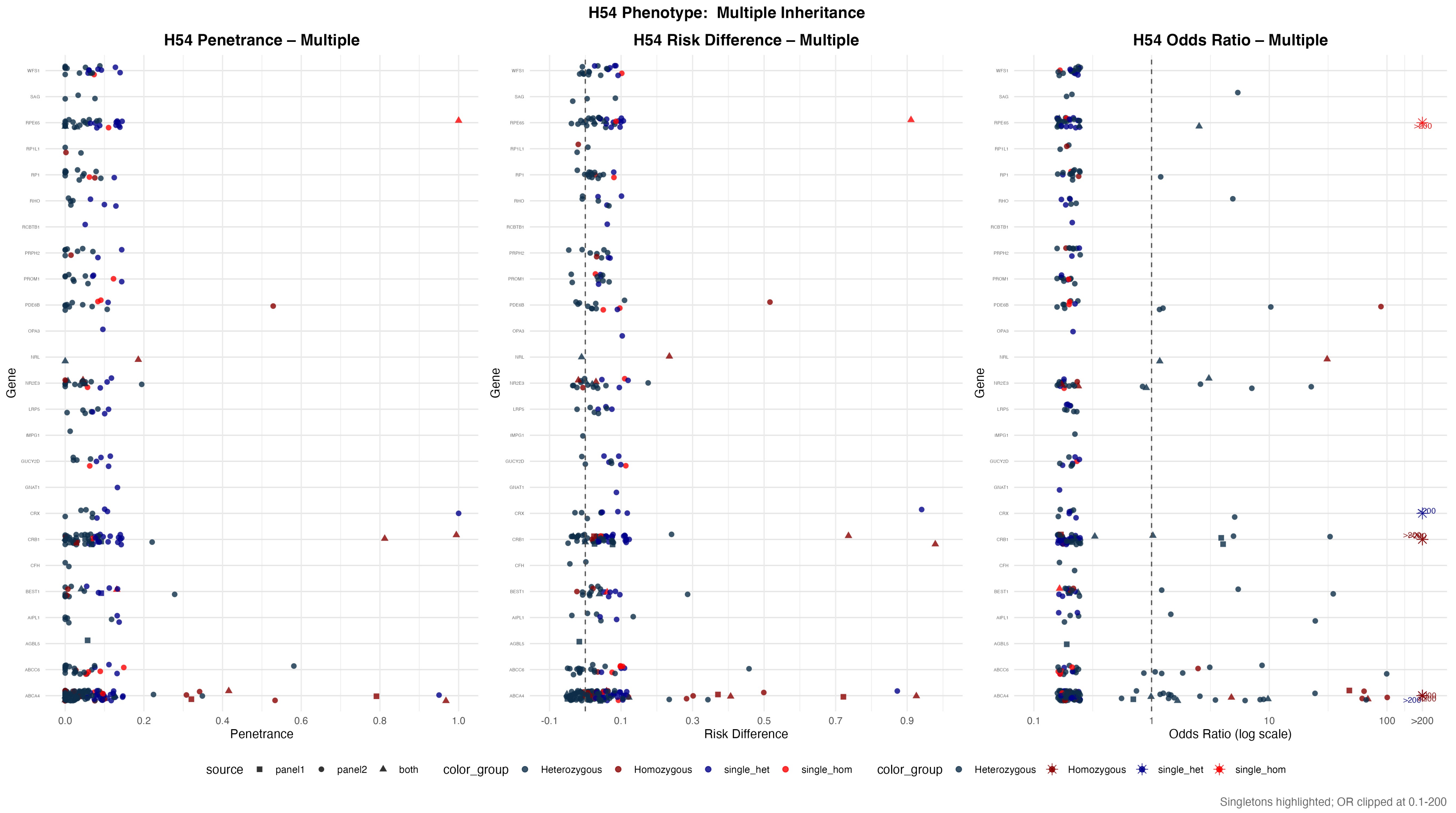
H54 Phenotype: Multiple Inheritance
H54 Penetrance – Multiple
H54 Risk Difference – Multiple
H54 Odds Ratio – Multiple
Gene
WFS1
SAG
RPE65
RP1L1
RP1
RHO
RCBTB1
PRPH2
PROM1
PDE6B
OPA3
NRL
NR2E3
LRP5
IMPG1
GUCY2D
GNAT1
CRX
CRB1
CFH
BEST1
AIPL1
AGBL5
ABCC6
ABCA4
Penetrance
Risk Difference
Odds Ratio (log scale)
source
panel1
panel2
both
color_group
Heterozygous
Homozygous
single_het
single_hom
>200
Singletons highlighted; OR clipped at 0.1-200

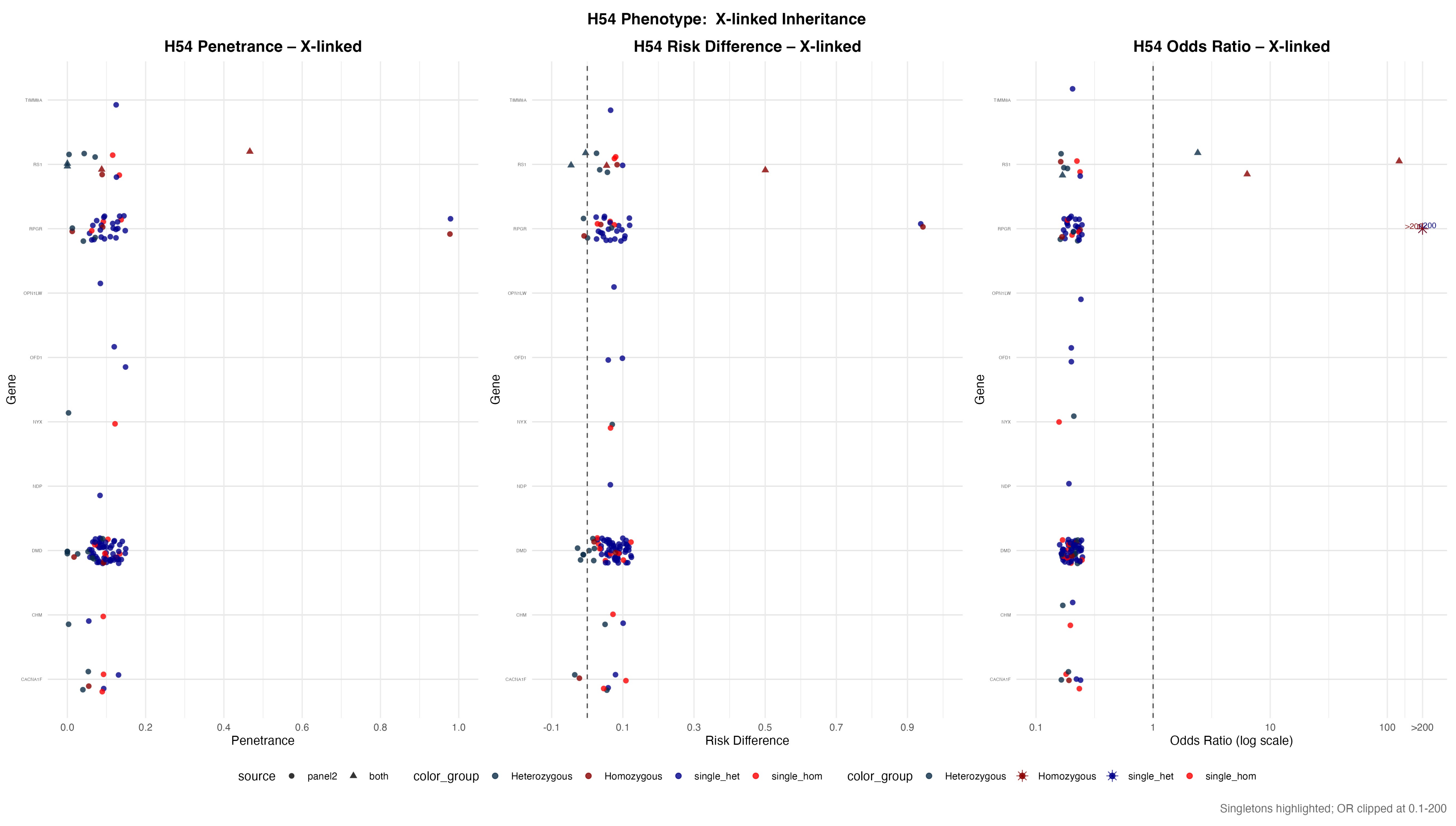

H54 Phenotype: X-linked Inheritance
H54 Penetrance – X-linked
H54 Risk Difference – X-linked
H54 Odds Ratio – X-linked
Gene
TIMM8A
RS1
RPGR
OPN1LW
OFD1
NYX
NDP
DMD
CHM
CACNA1F
0.0
0.2
0.4
0.6
0.8
1.0
Penetrance
-0.1
0.1
0.3
0.5
0.7
0.9
Risk Difference
0.1
1
10
100
>200
Odds Ratio (log scale)
>200
>200
source
panel2
both
color_group
Heterozygous
Homozygous
single_het
single_hom
Singletons highlighted; OR clipped at 0.1-200

**Supplementary Figure 9. Variant-Level Penetrance and Association Metrics for Retinal Disorders (H54 Subchapter).**

Estimated clinical penetrance (left), risk difference (center), and odds ratio (right) are shown for heterozygous and homozygous carriers of known IRD-associated variants, aggregated by gene and stratified by inheritance mode: (A) autosomal recessive (AR), (B) autosomal dominant (AD), (C) multiple inheritance patterns, and (D) X-linked genes. Each point represents a variant with linked genotype and phenotype data from 81,080 carriers. Outcomes are defined as presence of at least one ICD-10 code within subchapter H54 (visual impairment including blindness). Colors and shapes denote zygosity and variant source (Panel I vs. Panel II). Vertical dashed lines indicate the null values for risk difference (0) and odds ratio (1). Variants with extreme OR values are plotted at axis limits for clarity. This subchapter captures advanced stages of retinal degeneration, enabling quantification of variant expressivity in the context of severe vision loss.

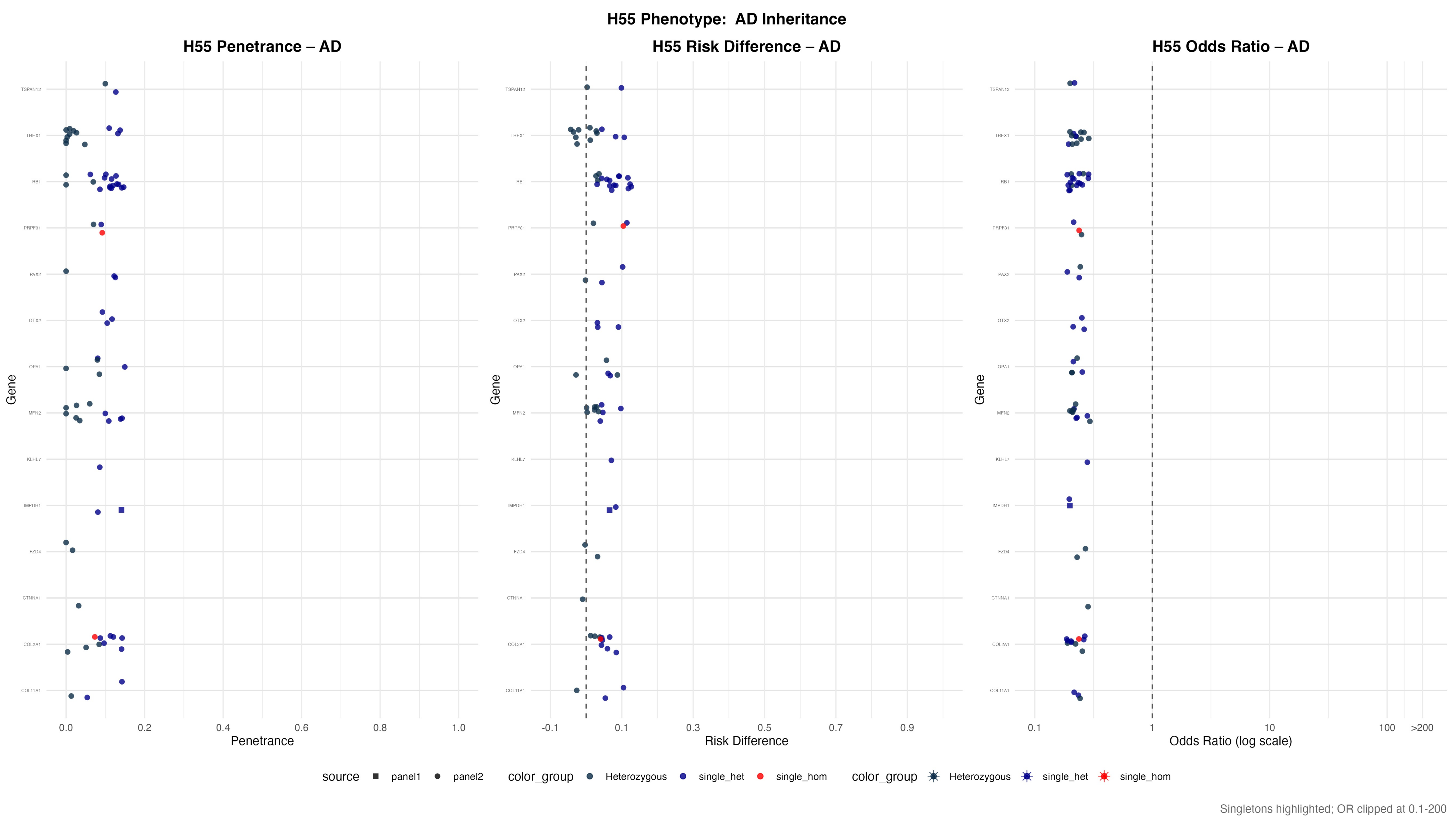
H55 Phenotype: AD Inheritance
H55 Penetrance – AD
H55 Risk Difference – AD
H55 Odds Ratio – AD
Gene
TSPAN12
TREX1
RB1
PRPF31
PAX2
OTX2
OPA1
MFN2
KLHL7
IMPDH1
FZD4
CTNNA1
COL2A1
COL11A1
0.0
0.2
0.4
0.6
0.8
1.0
Penetrance
-0.1
0.1
0.3
0.5
0.7
0.9
Risk Difference
0.1
1
10
100
>200
Odds Ratio (log scale)
source
panel1
panel2
color_group
Heterozygous
single_het
single_hom
Singletons highlighted; OR clipped at 0.1-200

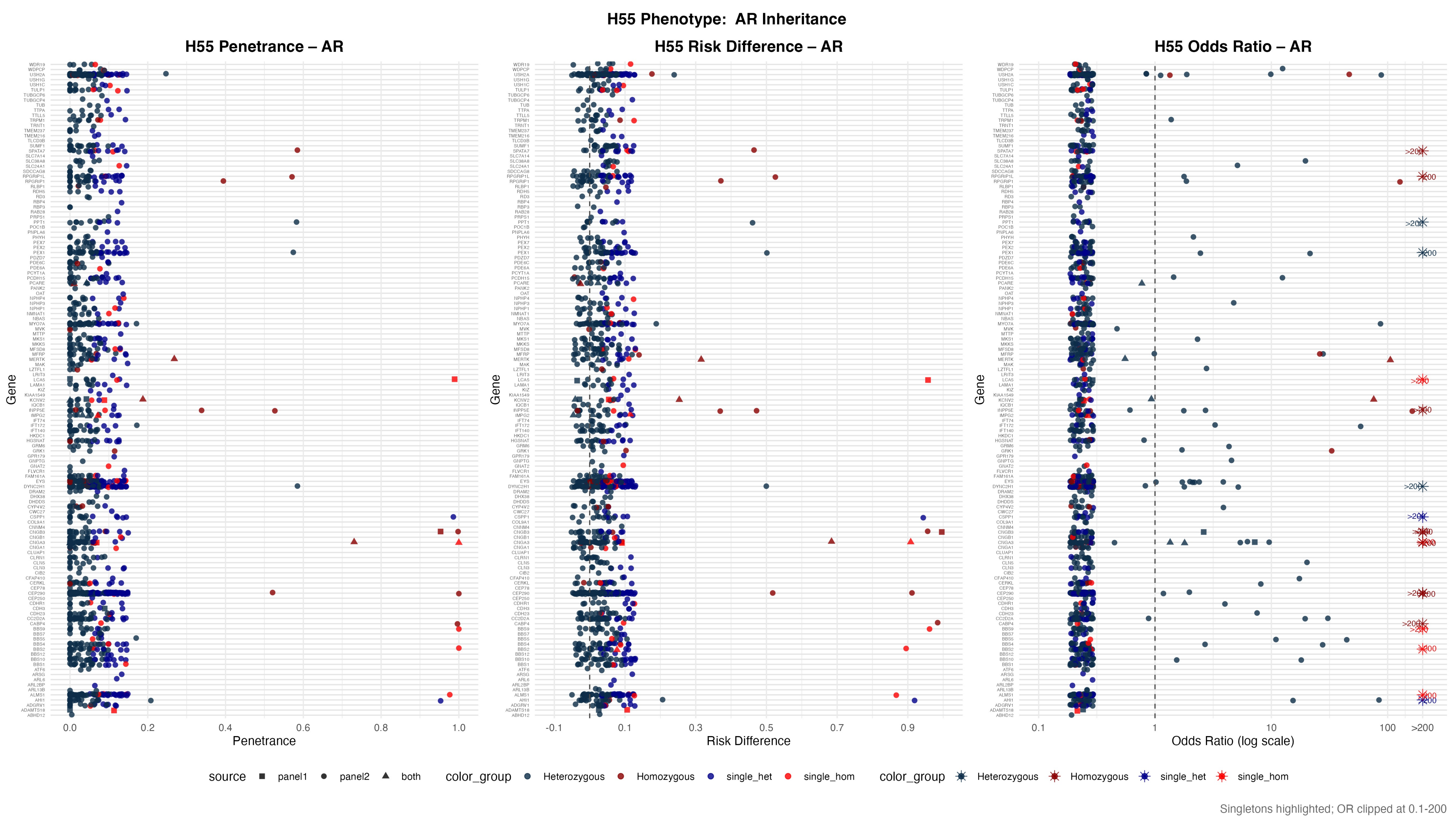
H55 Phenotype: AR Inheritance
H55 Penetrance – AR
H55 Risk Difference – AR
H55 Odds Ratio – AR
Gene
Penetrance
Risk Difference
Odds Ratio (log scale)
source
panel1
panel2
both
color_group
Heterozygous
Homozygous
single_het
single_hom
Singletons highlighted; OR clipped at 0.1-200

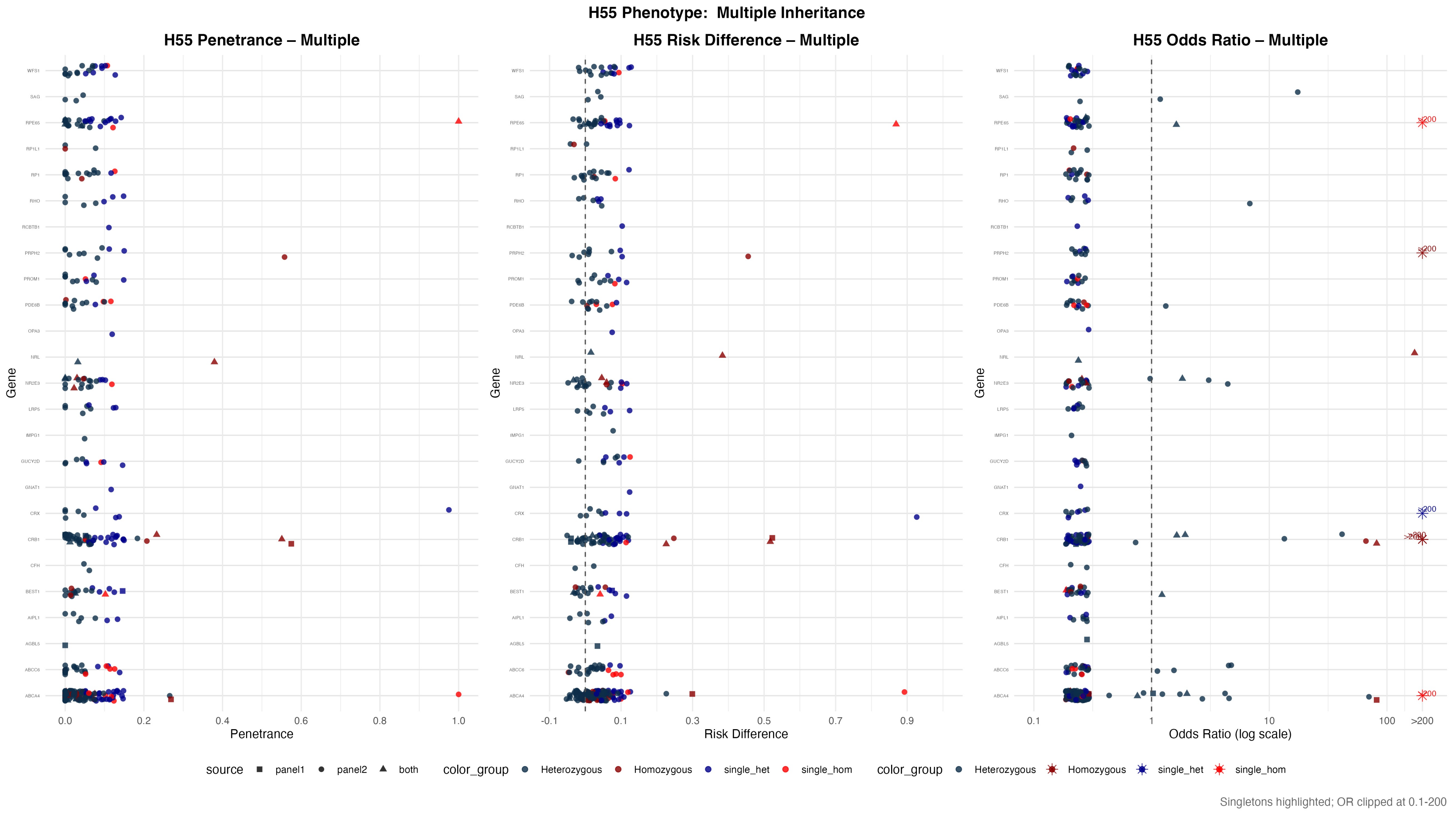

H55 Phenotype: Multiple Inheritance
H55 Penetrance – Multiple
H55 Risk Difference – Multiple
H55 Odds Ratio – Multiple
Gene
Penetrance
Risk Difference
Odds Ratio (log scale)
WFS1
SAG
RPE65
RP1L1
RP1
RHO
RCBTB1
PRPH2
PROM1
PDE6B
OPA3
NRL
NR2E3
LRP5
IMPG1
GUCY2D
GNAT1
CRX
CRB1
CFH
BEST1
AIPL1
AGBL5
ABCC6
ABCA4
source
panel1
panel2
both
color_group
Heterozygous
Homozygous
single_het
single_hom
>200
Singletons highlighted; OR clipped at 0.1-200

### H55 Phenotype: X-linked Inheritance

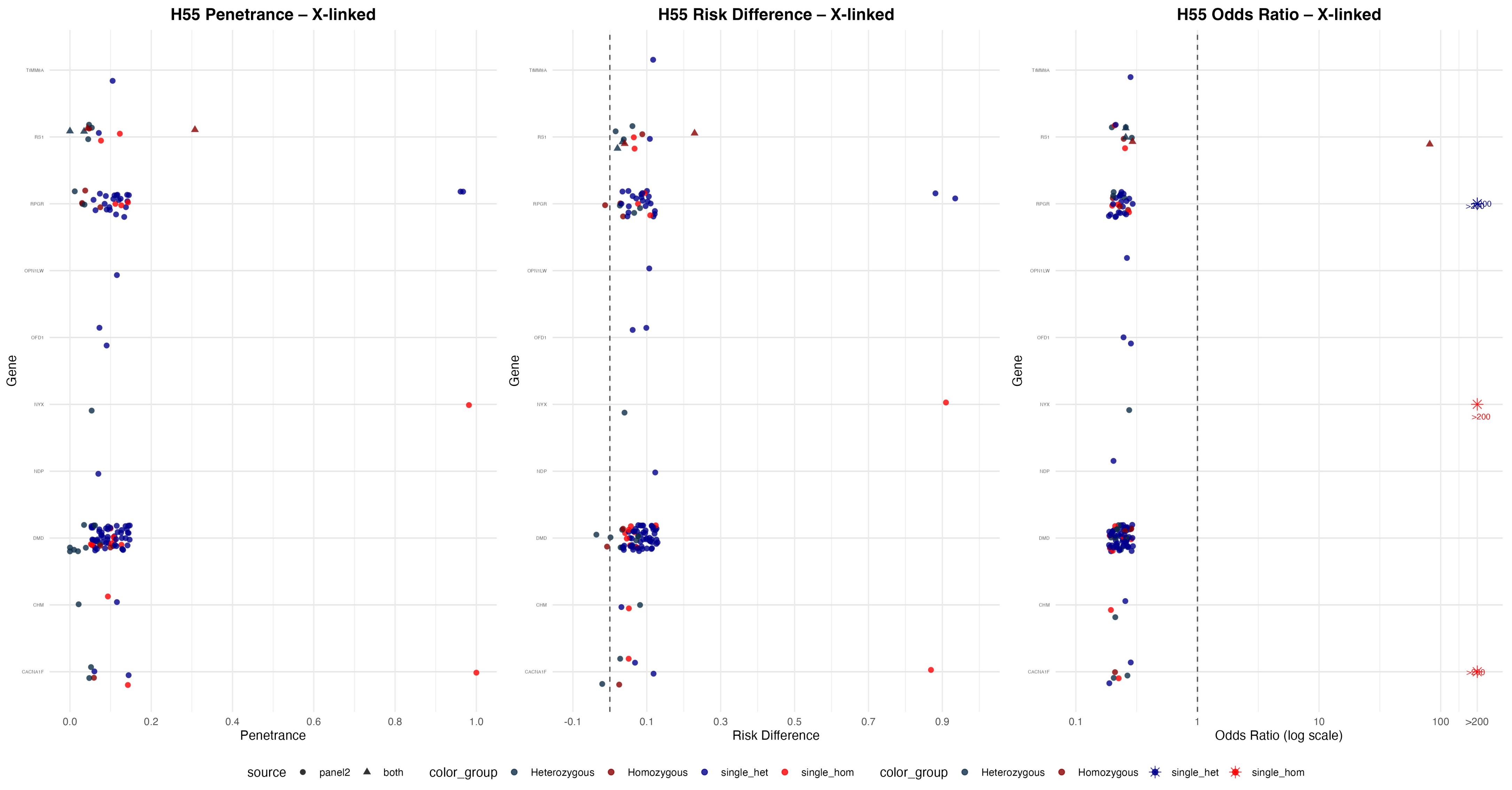


Singletons highlighted; OR clipped at 0.1-200

**Supplementary Figure 10. Variant-Level Penetrance and Association Metrics Across Inheritance Modes (H55 Subchapter).**

Estimated clinical penetrance (left), risk difference (center), and odds ratio (right) are shown for heterozygous and homozygous carriers of known IRD-associated variants, aggregated by gene and stratified by inheritance mode: (A) autosomal recessive (AR), (B) autosomal dominant (AD), (C) multiple inheritance patterns, and (D) X-linked genes. Each point represents a variant with linked genotype and phenotype data from 81,080 carriers. Outcomes are defined as presence of at least one ICD-10 code within subchapter H55 (nystagmus and other irregular eye movements). Colors and shapes denote zygosity and variant source (Panel I vs. Panel II). Vertical dashed lines indicate the null values for risk difference (0) and odds ratio (1). Variants with extreme OR values are plotted at axis limits for clarity. This subchapter-specific analysis captures early neuro-ophthalmic manifestations commonly seen in IRD, including involuntary eye movements and related visual instability.

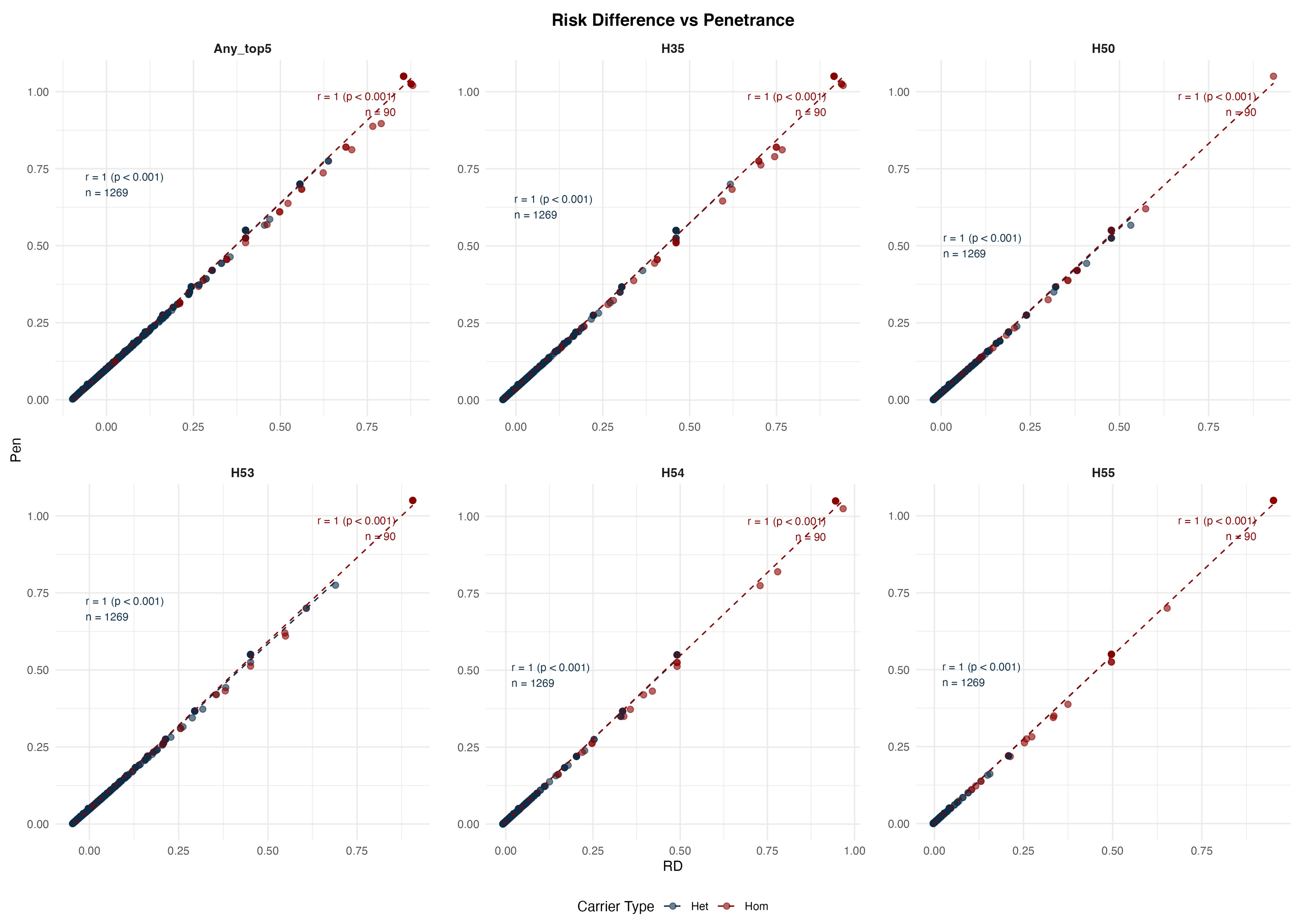
Risk Difference vs Penetrance
Any_top5
H35
H50
H53
H54
H55
r = 1 (p < 0.001)
n = 1269
r = 1 (p < 0.001)
n = 90
Pen
RD
Carrier Type
Het
Hom

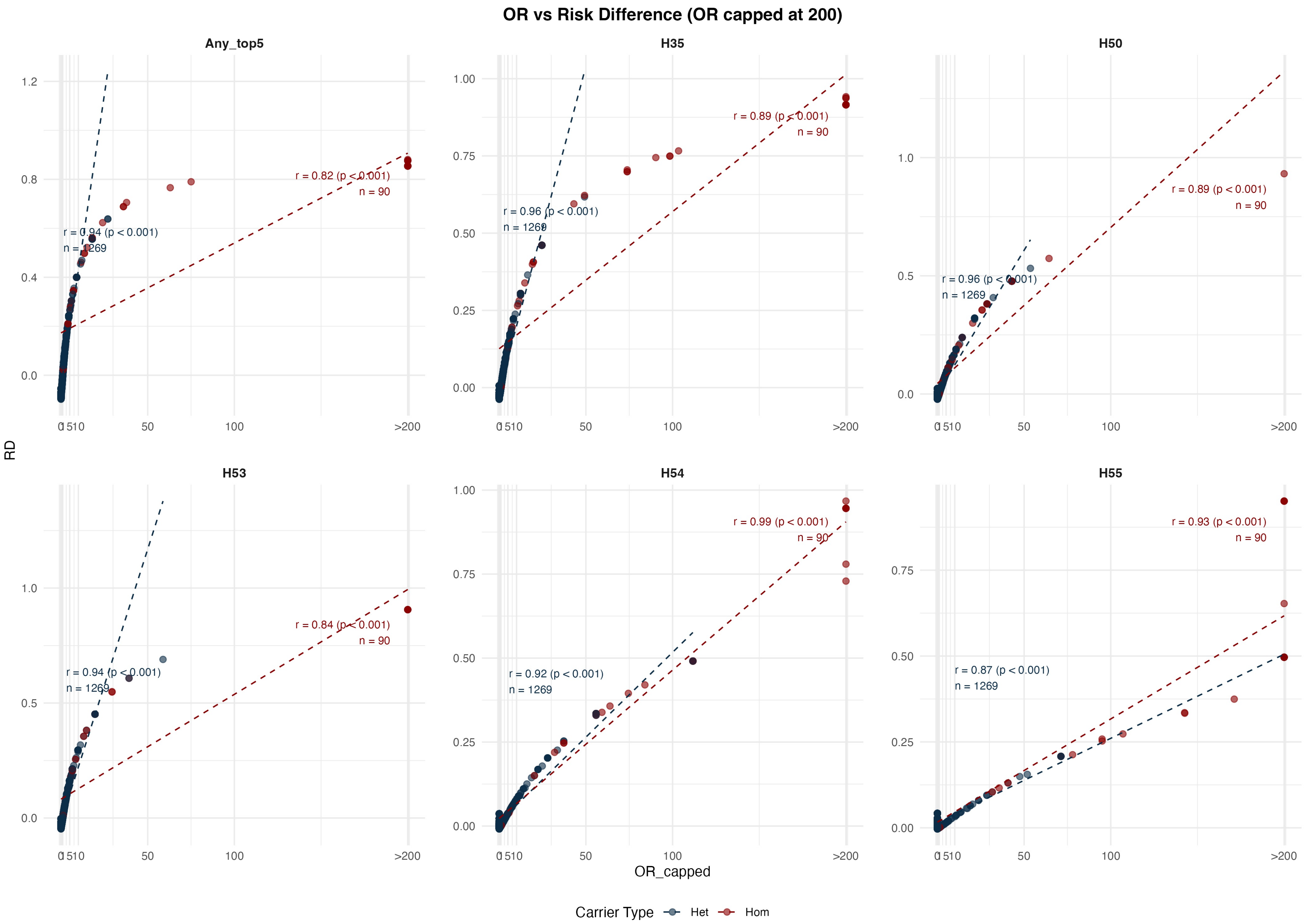
OR vs Risk Difference (OR capped at 200)
Any_top5
H35
H50
H53
H54
H55
r = 0.94 (p < 0.001)
n = 1269
r = 0.82 (p < 0.001)
n = 90
r = 0.96 (p < 0.001)
n = 1269
r = 0.89 (p < 0.001)
n = 90
r = 0.96 (p < 0.001)
n = 1269
r = 0.89 (p < 0.001)
n = 90
r = 0.94 (p < 0.001)
n = 1269
r = 0.84 (p < 0.001)
n = 90
r = 0.92 (p < 0.001)
n = 1269
r = 0.99 (p < 0.001)
n = 90
r = 0.87 (p < 0.001)
n = 1269
r = 0.93 (p < 0.001)
n = 90
RD
OR_capped
Carrier Type
Het
Hom

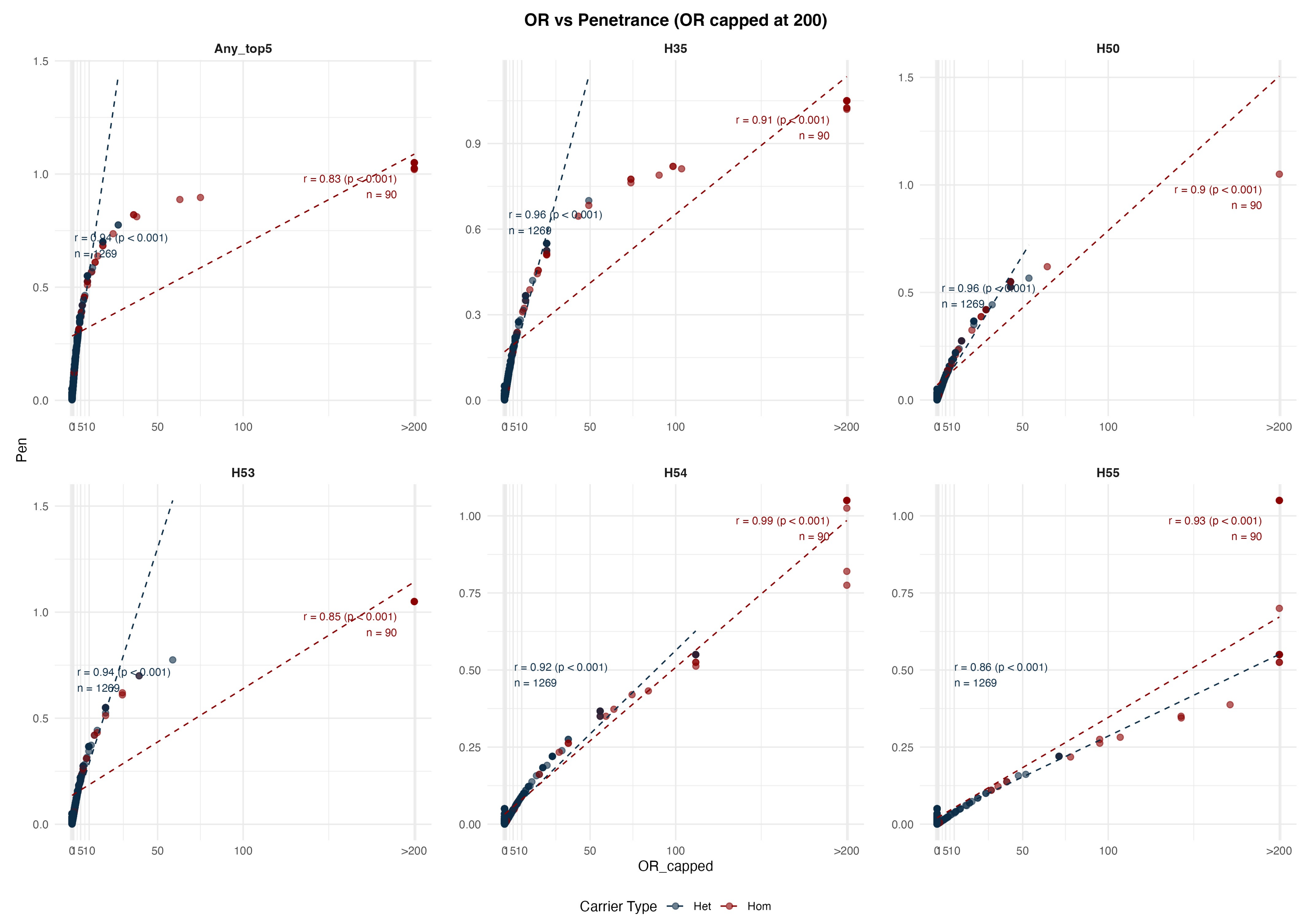
OR vs Penetrance (OR capped at 200)
Any_top5
r = 0.94 (p < 0.001)
n = 1269
r = 0.83 (p < 0.001)
n = 90
H35
r = 0.96 (p < 0.001)
n = 1269
r = 0.91 (p < 0.001)
n = 90
H50
r = 0.96 (p < 0.001)
n = 1269
r = 0.9 (p < 0.001)
n = 90
H53
r = 0.94 (p < 0.001)
n = 1269
r = 0.85 (p < 0.001)
n = 90
H54
r = 0.92 (p < 0.001)
n = 1269
r = 0.99 (p < 0.001)
n = 90
H55
r = 0.86 (p < 0.001)
n = 1269
r = 0.93 (p < 0.001)
n = 90
Pen
OR_capped
Carrier Type
Het
Hom

**Supplementary Figure 11. Correlations between variant-level penetrance, odds ratio (OR), and risk difference (RD) across ICD-10 subchapters.**

Each panel shows scatterplots of pairwise correlations between penetrance, odds ratio (OR, capped at 200), and risk difference (RD), computed separately for heterozygous (blue) and homozygous (dark red) carriers. Metrics were derived from variant-level associations across six ICD-10 subchapter groupings: H35, H50, H53, H54, H55, and the composite outcome Any_top5, which aggregates all five. Variants included had more than one carrier of the given zygosity. Pearson correlation coefficients (r), p-values, and sample sizes (n) are annotated within each facet. Dashed red lines represent linear regression fits. These analyses confirm strong positive relationships between penetrance and both OR and RD, and between OR and RD themselves, across IRD-associated variants. While OR and RD are both measures of effect size, penetrance provides a more interpretable measure of expressivity within the cohort.

Pedigrees for two unrelated families carrying the KCNV2 variant (chr9:2718166:G>T; c.427G>T) can be made available upon reasonable request from the corresponding author.

**Supplementary Figure 12. Variable Penetrance of Highly Penetrant KCNV2 Variant (chr9:2718166:G>T; c.427G>T) Across Two Unrelated Families.**

Pedigrees illustrate two unrelated families carrying the same KCNV2 variant with differing penetrance patterns. Ages are shown within each symbol, and values underneath indicate the individual's inbreeding coefficient (plink --het). Phenotype status is based on presence of any of the top five IRD-related ICD-10 phenotypes (Top5 phenotypes: H35, H54, H55, H50, H53). (A) Family showing 50% penetrance in homozygous carriers and 100% penetrance in heterozygous carriers. (B) Family showing 50% penetrance in homozygous carriers and 0% penetrance in heterozygous carriers. Color coding: white = non-carrier, healthy; dark red = homozygous, affected with top five phenotypes; red = homozygous, healthy; grey = heterozygous, healthy; dark blue = heterozygous, affected with top five phenotypes. Question marks denote individuals not included in the EGP.

Pedigree for the family carrying the CYP27A1 variant (chr2:218814188:G>A) can be made available upon reasonable request from the corresponding author.

**Supplementary Figure 13. Pedigree of a family segregating pathogenic variants in CYP27A1 and ABCA4.**

This family includes a male with dual findings: homozygous for the CYP27A1 variant (chr2:218814188:G>A; P; two stars) and heterozygous for the ABCA4 variant (chr1:94008251:C>T; P/LP; two stars). He presents with a combination of vision-related (H57.9: unspecified disorder of eye and adnexa) and neurodevelopmental diagnoses (autism and disorders of learning), consistent with cerebrotendinous xanthomatosis (CTX), a treatable autosomal recessive disorder known to cause early visual dysfunction. Multiple family members are heterozygous for one or both variants. Of note, a female in her thirties (rightmost individual) heterozygous for both CYP27A1 and ABCA4 carries an H53.8 diagnosis (visual disturbance), suggesting possible modifier effects or partial penetrance. (A) Pedigree annotated for the CYP27A1 variant, showing the homozygous proband and heterozygous relatives. (B) Pedigree annotated for the ABCA4 variant, showing heterozygous carriers including the affected female with one of the top five phenotypes (H53). Color coding: dark red = homozygous affected; dark blue = heterozygous affected; grey = heterozygous healthy; white = non-carrier healthy. Ages are shown within each symbol, and values underneath indicate inbreeding coefficients (plink --het). Question marks denote individuals not sequenced in the EGP.
